# Dietary patterns for weight and glycemic management in persons with type 1 diabetes: a meta-analysis of clinical trials

**DOI:** 10.1101/2025.05.23.25328241

**Authors:** Daria Igudesman, Laura M. Nally, Alyssa A. Grimshaw, Craig Gunderson, Elizabeth G. Considine, Laura M. Jacobsen, Mustafa Tosur, Peter A. Gottlieb, Irl Hirsch, Lori Laffel, Jennifer L. Sherr, Chantal Mathieu, Richard E. Pratley

**Author notes:** Corresponding Authors: Daria Igudesman, PhD, RD 301 E Princeton St., Orlando, FL 32804 Laura Nally, MD 1 Long Wharf Drive, Suite 503, New Haven, CT 06511. These authors contributed equally (Daria Igudesman, Laura M. Nally). **PROSPERO Registration:** CRD42024519941.

## Abstract

**Background:** Medical nutrition therapy is fundamental for managing glycemia and weight in type 1 diabetes, yet dietary guidance specific to this population and relevant subgroups is lacking.

**Purpose:** We synthesized the interventional literature investigating diet patterns for glycemic and weight management in youth and adults with type 1 diabetes, with attention to interindividual variation that suggests the need for precision approaches.

**Data Sources:** AMED, CINAHL, Cochrane Library, Ovid MEDLINE, Ovid Embase, Google Scholar, and Web of Science Core Collection were searched from January 2011 to June 2024.

**Study Selection:** Clinical trials ≥4 weeks with ≥10 youth and/or adults diagnosed with type 1 diabetes ≥6 months prior and reported HbA1c or weight were included.

**Data Synthesis:** Twelve studies with 668 participants were included. Data were pooled by random-effects models for HbA1c and weight. Studies with insufficient data and subgroup differences were narratively synthesized per Synthesis Without Meta-analysis guidelines. Pooled results of very low to moderate certainty evidence showed no advantage of any particular diet pattern relative to routine care in randomized trials. Very low-quality evidence from single-arm low carbohydrate trials suggested improved HbA1c over time (-0.63% [95%CI –0.99, -0.27]; -6.0 mmol/mol [-10.8, -3.0]). Wide pooled confidence intervals demonstrated between-person heterogeneity; however, stratification of results by participant characteristics was rarely performed.

**Limitations:** Limited evidence precluded subgroup analyses that can inform precision nutrition approaches.

**Conclusions:** Randomized trials designed to investigate treatment response heterogeneity are needed to determine whether precision nutrition therapies optimize glycemia and weight in persons with type 1 diabetes.

**Disclosure Summary:** Laura Nally receives research support from the National Institutes of Health and is a consultant for Medtronic, WebMD, and Calm. Jennifer Sherr serves, or has served, on advisory panels for Bigfoot Biomedical, Cecelia Health, Insulet Corporation, Mannkind, Medtronic Diabetes, StartUp Health Diabetes Moonshot, and Vertex. Dr. Sherr has served as a consultant to Abbott Diabetes, Bigfoot Biomedical, Insulet, Medtronic Diabetes, and Zealand. Yale School of Medicine has received research support for Dr. Sherr from Abbott Diabetes, JAEB Center for Health Research, JDRF, Insulet, Medtronic, NIH, and Prevention Bio. Chantal Mathieu serves or has served on the advisory panel for Novo Nordisk, Sanofi, Eli Lilly and Company, Novartis, Dexcom, Boehringer Ingelheim, Bayer, Roche, Abbott, Medtronic, Insulet, Biomea Fusion, SAB Bio and Vertex. Financial compensation for these activities has been received by KU Leuven; KU Leuven has received research support for Chantal Mathieu from Medtronic, Novo Nordisk, and Sanofi; Chantal Mathieu serves or has served on the speakers bureau for Novo Nordisk, Sanofi, Eli Lilly and Company, Medtronic, Dexcom, Insulet, Abbott, Vertex and Boehringer Ingelheim. Financial compensation for these activities has been received by KU Leuven. Irl Hirsch reports research funding from Tandem and Dexcom; and consulting fees from Abbott, Roche, GWave, and Vertex. Lori Laffel reports consulting for Dexcom, Boehringer Ingelheim, Medtronic, Provention Bio, Sanofi, Medtronic, Sequel MedTech, Vertex, and Tandem Diabetes. Peter Gottlieb has served as an advisor to Viacyte/Vertex, Imcyse, JDRF T1D Fund, and GentiBio; has received research support from Novo Nordisk, Imcyse, Novartis, Mercia/Nova, Provention Bio, ActoBio Therapeutics, Helmsley Foundation, JDRF, and NIH; and is a co-founder, Chief Medical Officer, and shareholder of ImmunoMolecular Therapeutics, Inc. Mustafa Tosur served as an advisory board member for Provention Bio in 2020 and 2021. Richard Pratley has received the following (thru 12/31/2023 directed to his institution; as of 1/1/2024 directed to Dr. Pratley personally): speaker fees from Lilly, Merck and Novo Nordisk; consulting fees from Bayer AG, Bayer HealthCare Pharmaceuticals, Inc., Corcept Therapeutics Incorporated, Dexcom, Endogenex, Inc., Gasherbrum Bio, Inc., Genprex, Getz Pharma, Hanmi Pharmaceutical Co., Hengrui (USA) Ltd., Intas Pharmaceuticals, Inc., Lilly, Merck, Novo Nordisk, Pfizer, Rivus Pharmaceuticals Inc., Sanofi, and Sun Pharmaceutical Industries; and grants from Biomea Fusion, Carmot Therapeutics, Dompe, Endogenex, Inc., Fractyl, Lilly, Novo Nordisk, and Sanofi. All other authors report no conflict of interest.

---

The prevalence of macrovascular disease in youth and adults with type 1 diabetes is markedly elevated relative to individuals without diabetes (1). Mitigation of this risk is contingent upon the attainment of glycemic targets in tandem with the prevention of excess weight gain (2) and traditional risk factors such as hypertension. Despite considerable advances in the development of diabetes technologies, a small proportion of youth and adults with type 1 diabetes meet glycemic and weight targets (3-5). Although medical nutrition therapy is a cornerstone of glycemic and weight management, a dearth of literature investigating dietary patterns in persons with type 1 diabetes (6) poses a barrier to achieving clinical targets. Without robust scientific evidence, clinicians may feel ill-equipped to recommend diet approaches to persons with type 1 diabetes (7).

Emerging findings from precision medicine highlight the possibility of augmenting treatment success by tailoring therapies to population subgroups, rather than recommending a one-size-fits-all approach (8, 9). In 2018, the Precision Medicine in Diabetes Initiative was launched by the American Diabetes Association (ADA) and European Association for the Study of Diabetes, with the first consensus report published in 2020 (10). For the type 1 diabetes treatment group, initial efforts focused on the impact of diabetes technologies (11); however, several other areas of interest were identified, with medical nutrition therapy being a top priority. This aligned with a recent focus of the National Institutes of Health (12) on precision nutrition, defined as the stratification of dietary recommendations by population subgroups (8, 13). The most recent systematic review of diet patterns for type 1 and type 2 diabetes management included studies published through the year 2010 and included both observational and experimental evidence (14). Rapid advances in diabetes technology over the past decade have likely augmented the safety and feasibility of diet patterns on diabetes management, potentially limiting the generalizability of earlier research to contemporary populations. Accordingly, the objective of this systematic review and meta-analysis was to synthesize data from clinical trials published between January 2011 and June 2024 that investigated the effects of diet patterns on glycemia and/or weight in youth and adults with type 1 diabetes, with particular attention to subgroup analyses that indicate the need to tailor treatments according to individual characteristics.

## Research Design and Methods

### Protocol Development, Data Sources and Searches

We performed a literature search in accordance with Cochrane recommendations with Preferred Reporting Items for Systematic Reviews and Meta-Analyses 2020 (**Supplemental Table 1, Supplemental Table 2**) (15) and prospectively registered the study protocol with PROSPERO (CRD42024519941, https://www.crd.york.ac.uk/prospero/display_record.php?ID=CRD42024519941). Literat ure searches were conducted in AMED, CINAHL, Cochrane Library, Ovid MEDLINE, Ovid Embase, Google Scholar, and Web of Science Core Collection from January 2011 to June 2024, to identify relevant published literature related to dietary patterns and type 1 diabetes. References of identified studies were manually searched. **Supplemental Table 3** shows full details of the search strategy. Forward and backward citation chasing was performed using CitationChaser.

### Study Selection

We included randomized and single-arm clinical trials ≥4 weeks that enrolled participants with type 1 diabetes aged ≥2 years with a diabetes duration of ≥6 months. Trials needed to include ≥10 participants per intervention arm, or a total of 10 participants for crossover trials. One of the co-primary review outcomes of hemoglobin A1c (HbA1c) or weight must have been reported. Full inclusion and exclusion criteria are listed in **Supplemental Table 4**.

### Abstract and Full-Text Screening

Two out of three reviewers (DI, LMN, and AAG) independently screened articles in a blinded fashion via the Covidence platform. Reviewers met to resolve disagreements.

### Data Extraction

EC and DI independently extracted data, which was verified by another reviewer. A standardized template was used for data extraction, which included information about the publication, study design, population, intervention, duration, and outcomes.

### Risk of Bias (Quality) Assessment

The Cochrane Risk of Bias 2 (16) tool was used to evaluate the risk of bias for randomized trials in five domains. The Risk of Bias in Non-randomised Studies - of Interventions (17) tool was applied to single-arm studies in seven domains. Results were visualized using the robvis tool (https://www.riskofbias.info/welcome/robvis-visualization-tool).

### GRADE Evidence Rating

In accordance with Cochrane guidelines (18), the GRADE approach was used to rate the overall quality of evidence for all studies, including those that could not be included in meta-analyses. Studies were grouped by study design (randomized or single-arm) and diet pattern (general healthful eating, low carbohydrate, Mediterranean, and low fat). Studies were assessed based on risk of bias, inconsistency, indirectness, imprecision, and publication bias. Randomized studies started as high certainty, whereas single-arm studies started as low certainty due to inherent confounding bias (19). Each body of evidence received a final grade of high, moderate, low, or very low certainty. DI and LMN independently conducted the GRADE assessments for all included studies and met to reach consensus. LJ and CG served as adjudicators.

### Data Synthesis and Analysis

For the outcomes of HbA1c and weight, we performed meta-analyses using the restricted maximum likelihood (20) to calculate pooled estimates and 95% confidence intervals (95%CIs), stratified by diet pattern and study design. For randomized controlled trials, we compared mean changes in outcomes between the active treatment and control group(s). Results are reported separately for each active treatment-control comparison, including for one study that compared a Mediterranean and a low fat diet pattern to a low carbohydrate comparator (21). For single-arm non-randomized studies, baseline was compared to post-intervention. When means or standard deviations were not reported, we estimated the mean from the median and standard deviation from standard errors or CIs using guidance from the Cochrane Handbook (22). Statistical heterogeneity across studies was assessed using the I^2^ statistic and Cochrane’s Q-statistic (23). All statistical analyses, including the generation of forest plots displaying pooled results, were performed using Stata/BE, version 18.5 (StataCorp, College Station, TX).

When meta-analysis could not be performed due to insufficient data, narrative synthesis was employed following Synthesis Without Meta-analysis guidelines (24) (**Supplemental Table 5**). To assess the certainty of narratively synthesized findings, measures of dispersion reported by individual study authors are described in text alongside mean treatment effects, as available. Given the explicit aim to inform future precision nutrition approaches for optimizing health outcomes in persons with type 1 diabetes, we narratively synthesized heterogeneity in intervention effects within sociodemographic and clinical strata or in association with participant characteristics, when reported. Due to the interdependency of changes in weight on insulin dose, we narratively synthesized results for insulin dose.

## Results

### Searches

Our search yielded 5,131 records (**Supplemental Figure 1**). After duplicates were removed and titles and abstracts screened (**Supplemental Table 6**), 53 studies were assessed for eligibility during full-text evaluation. Twelve studies that enrolled 668 participants as part of 11 unique clinical trials—seven randomized and four single-arm— met pre-specified criteria for inclusion (**Table 1**). One trial reported the results of two study arms separately (25, 26). All studies reported HbA1c. Eight studies, including five randomized trials (21, 27-33), reported weight. Studies enrolled participants from ages 2 to 70 and spanned seven countries.

**Table 1.**
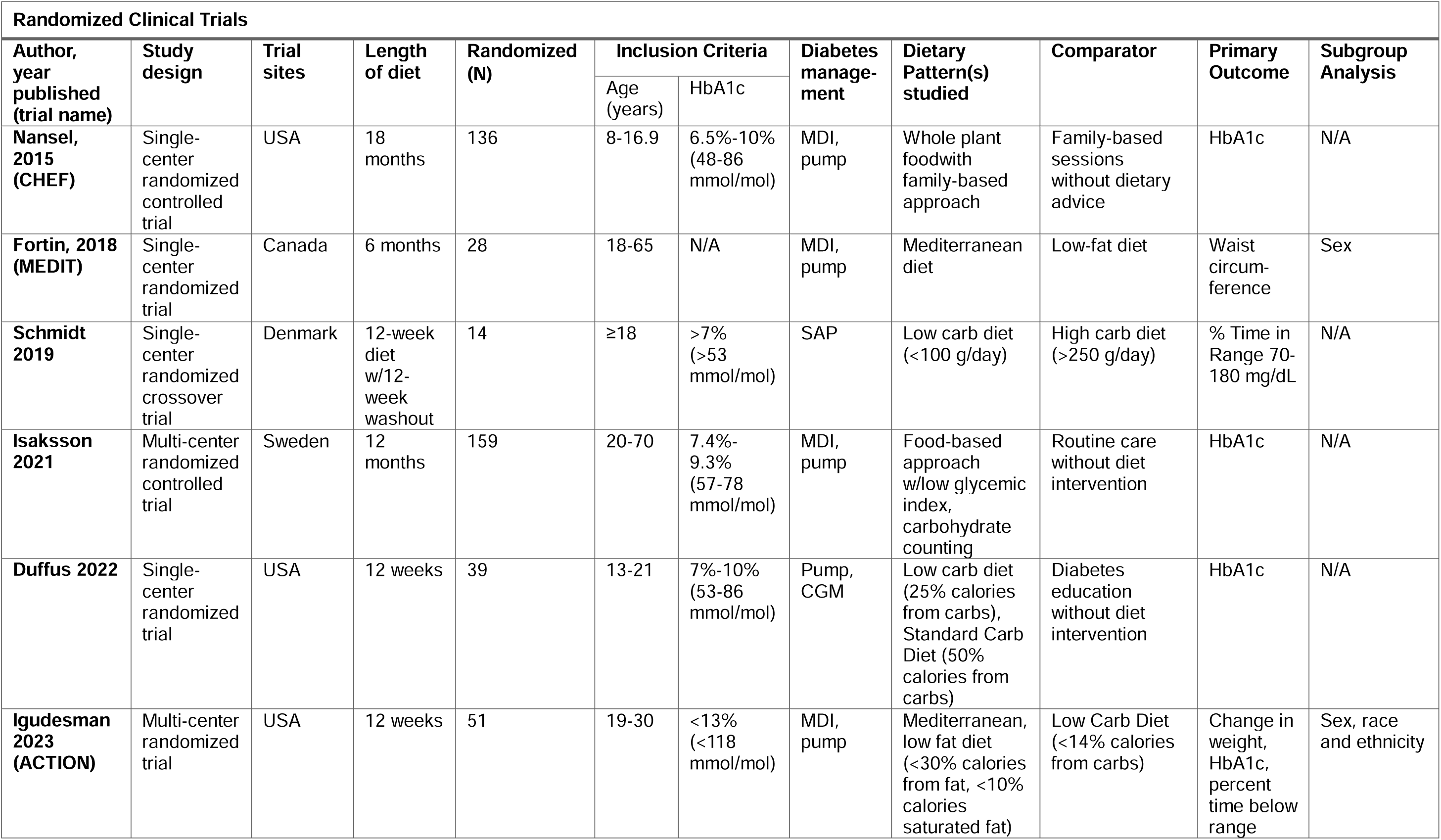
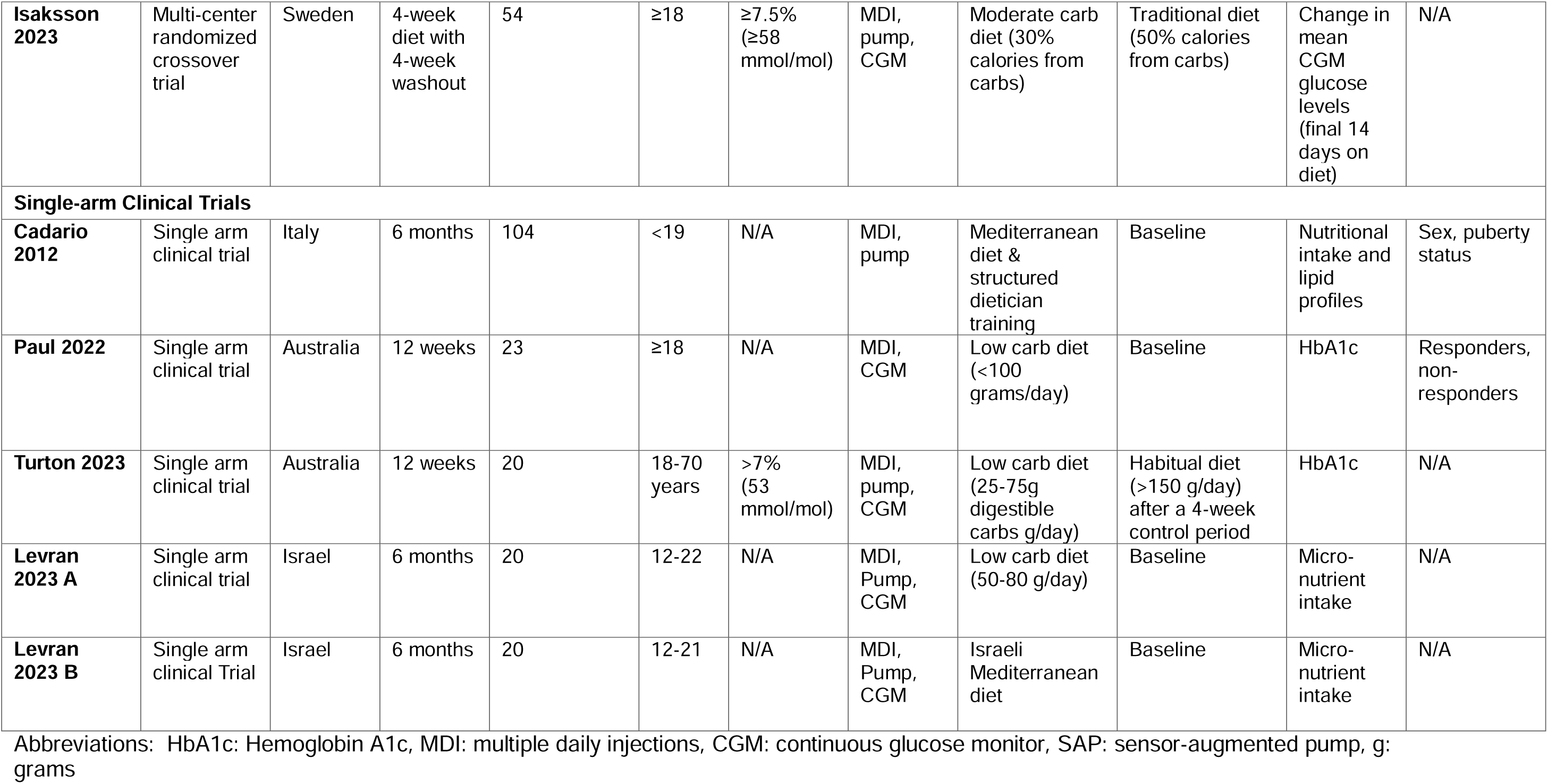
Clinical trials of dietary patterns included in the systematic review.

| Randomized Clinical Trials |  |  |  |  |  |  |  |  |  |  |  |
| --- | --- | --- | --- | --- | --- | --- | --- | --- | --- | --- | --- |
| Author, year published (trial name) | Study design | Trial sites | Length of diet | Randomized (N) | Inclusion Criteria |  | Diabetes management | Dietary Pattern(s) studied | Comparator | Primary Outcome | Subgroup Analysis |
|  |  |  |  |  | Age (years) | HbA1c |  |  |  |  |  |
| Nansel, 2015 (CHEF) | Single-center randomized controlled trial | USA | 18 months | 136 | 8-16.9 | 6.5%-10% (48-86 mmol/mol) | MDI, pump | Whole plant foodwith family-based approach | Family-based sessions without dietary advice | HbA1c | N/A |
| Fortin, 2018 (MEDIT) | Single-center randomized trial | Canada | 6 months | 28 | 18-65 | N/A | MDI, pump | Mediterranean diet | Low-fat diet | Waist circum-ference | Sex |
| Schmidt 2019 | Single-center randomized crossover trial | Denmark | 12-week diet w/12-week washout | 14 | ≥18 | >7% (>53 mmol/mol) | SAP | Low carb diet (<100 g/day) | High carb diet (>250 g/day) | % Time in Range 70-180 mg/dL | N/A |
| Isaksson 2021 | Multi-center randomized controlled trial | Sweden | 12 months | 159 | 20-70 | 7.4%-9.3% (57-78 mmol/mol) | MDI, pump | Food-based approach w/low glycemic index, carbohydrate counting | Routine care without diet intervention | HbA1c | N/A |
| Duffus 2022 | Single-center randomized trial | USA | 12 weeks | 39 | 13-21 | 7%-10% (53-86 mmol/mol) | Pump, CGM | Low carb diet (25% calories from carbs), Standard Carb Diet (50% calories from carbs) | Diabetes education without diet intervention | HbA1c | N/A |
| Igudesman 2023 (ACTION) | Multi-center randomized trial | USA | 12 weeks | 51 | 19-30 | <13% (<118 mmol/mol) | MDI, pump | Mediterranean, low fat diet (<30% calories from fat, <10% calories saturated fat) | Low Carb Diet (<14% calories from carbs) | Change in weight, HbA1c, percent time below range | Sex, race and ethnicity |
| **Isaksson 2023** | Multi-center randomized crossover trial | Sweden | 4-week diet with 4-week washout | 54 | ≥18 | ≥7.5% (≥58 mmol/mol) | MDI, pump, CGM | Moderate carb diet (30% calories from carbs) | Traditional diet (50% calories from carbs) | Change in mean CGM glucose levels (final 14 days on diet) | N/A |
| **Single-arm Clinical Trials** |  |  |  |  |  |  |  |  |  |  |  |
| **Cadario 2012** | Single arm clinical trial | Italy | 6 months | 104 | <19 | N/A | MDI, pump | Mediterranean diet & structured dietician training | Baseline | Nutritional intake and lipid profiles | Sex, puberty status |
| **Paul 2022** | Single arm clinical trial | Australia | 12 weeks | 23 | ≥18 | N/A | MDI, CGM | Low carb diet (<100 grams/day) | Baseline | HbA1c | Responders, non-responders |
| **Turton 2023** | Single arm clinical trial | Australia | 12 weeks | 20 | 18-70 years | >7% (53 mmol/mol) | MDI, pump, CGM | Low carb diet (25-75g digestible carbs g/day) | Habitual diet (>150 g/day) after a 4-week control period | HbA1c | N/A |
| **Levrán 2023 A** | Single arm clinical trial | Israel | 6 months | 20 | 12-22 | N/A | MDI, Pump, CGM | Low carb diet (50-80 g/day) | Baseline | Micro-nutrient intake | N/A |
| **Levrán 2023 B** | Single arm clinical Trial | Israel | 6 months | 20 | 12-21 | N/A | MDI, Pump, CGM | Israeli Mediterranean diet | Baseline | Micro-nutrient intake | N/A |

Abbreviations: HbA1c: Hemoglobin A1c, MDI: multiple daily injections, CGM: continuous glucose monitor, SAP: sensor-augmented pump, g: grams

### Diet Patterns Identified

Among the 12 selected studies, there were six low or moderate carbohydrate interventions, four Mediterranean (including one Israeli Mediterranean), one low fat diet, and two healthful eating approaches (whole food plant-based, and food-based approach with low glycemic index). The moderate and low carbohydrate diet interventions involved limiting total carbohydrates to 25% (30) or 50% (34) of total caloric intake, less than 100 grams per day (28, 31) or a specific range of daily carbohydrate intake (50-80 grams per day (32) or 25-75 grams of digestible carbohydrates per day (25). Mediterranean diets encouraged use of olive oil, plant-based foods, whole grains, and fish, and limited red meat and processed foods. One study tested a Mediterranean diet and a hypocaloric low fat diet (<30% calories from fat and <10% calories from saturated fat) against a hypocaloric low carbohydrate diet (21). This study was the only one to explicitly recommend caloric reduction given the aim to promote weight loss in participants with overweight or obesity (21). The healthful diet patterns included a family-based behavioral intervention to increase intake of whole plant foods, including fruit, vegetables, whole grains, legumes, nuts, and seeds (29), and a food-based approach encouraging low glycemic index foods (35).

### Risk of Bias

Among the seven randomized trials reporting HbA1c, one was deemed to be at a low risk of bias (29), four had some concerns (21, 27, 30, 35), and two were judged to have a high risk of bias—one due to missing data (28) and the second due to potential carryover effects in a randomized crossover design (34) (**Supplemental Figure 2**). Of the randomized trials reporting weight, three had some concerns (21, 27, 35) and two had a high risk of bias due to missing data (28) and bias in the measurement of the outcome (28, 34). All non-randomized trials were found to be at an overall high risk of bias for both HbA1c (**Supplemental Figure 3**) and weight (**Supplemental Figure 4**), as none of the included trials adjusted for potential time-varying confounders in their pre-post analyses. With respect to reporting bias, selective non-publication was deemed to be unlikely given the large proportion of studies with null results. Similarly, none of the studies were deemed to have been likely to selectively report results.

### GRADE Evidence Rating

Rationale for downgrading in each of the GRADE domains is comprehensively outlined in **Supplemental Table 11** and the associated supplemental text. All bodies of evidence were downgraded by one point in the indirectness domain due to a lack of generalizability. Study participants tended to have lower mean HbA1c values and did not reflect the sociodemographic characteristics, such race and ethnicity, of the broader target population of individuals living with type 1 diabetes (3) (**Supplemental Table 12**). In contrast, no studies were downgraded for publication bias. All single-arm trials were deemed to have very serious limitations and thus downgraded by two levels of certainty due to a serious risk of bias in the confounding domain. Bodies of evidence graded as having moderate certainty included randomized trials of healthful diet patterns for the outcomes of HbA1c and weight, and randomized trials of low fat diets for the outcome of weight. Low certainty bodies of evidence included randomized trials of Mediterranean diet patterns for both HbA1c and weight, and randomized trials of low fat diet patterns for HbA1c. All other bodies of evidence, including all evidence from single-arm trials, were graded as very low certainty.

### Meta-Analyses and Narrative Syntheses

#### Impact of Dietary Patterns on Hemoglobin A1c

##### Healthful Diet Pattern

Two randomized trials investigated healthful diet patterns in youth (29) and adults (35) with type 1 diabetes, with the latter focusing on low glycemic index (35). Control arms included standard of care with carbohydrate counting (35) and usual care without a dietary intervention (29, 35). The pooled result from the meta-analysis of 136 youth and 133 adults was null, indicating no differential effect of healthful diets on HbA1c relative to control (–0.15% [95%CI –0.40, 0.11%]; -1.6 mmol/mol [95%CI –4.4, 1.2]; **Supplemental Table 7**).

##### Low Carbohydrate Diet Pattern

Six trials—three randomized (28, 30, 34) and three single-arm (25, 31, 32), studied the effects of low carbohydrate (15-25% calories from carbohydrates or 25 to <100 g/day) (25, 28, 30-32) or moderate carbohydrate (30% calories from carbohydrates) (34) diets on HbA1c in youth and adults with type 1 diabetes. Control groups included a standard carbohydrate diet (50% calories from carbohydrate) (30, 34), general diabetes education (30), and a high/traditional carbohydrate diet (>250 g/day or 50% calories from carbohydrate) (28, 34). Of the three randomized trials investigating low carbohydrate diets, two studies representing 56 participants with type 1 diabetes had available HbA1c data for meta-analysis (28, 30). Pooled results indicated no differential effects of a low carbohydrate diet on HbA1c relative to a high carbohydrate diet (28), a standard carbohydrate diet (30), or general diabetes education (30) over 4.4 months (0.32% [95%CI –0.07, 0.72%]; 3.5 mmol/mol [95%CI -0.8, 7.9]; **Figure 1A**). Due to insufficient data, the randomized crossover trial comparing a moderate carbohydrate diet to a traditional diet in 50 adults with type 1 diabetes was not included in pooled analysis but similarly found no difference in HbA1c change between study arms over 4.4 months (-0.10% [95%CI -0.25, 0.05%]; -1.1 mmol/mol [95%CI -2.7, 0.5]) (34). However, time in range (70-180 mg/dL) measured using continuous glucose monitoring (CGM) was nearly 5 percentage points higher during the final two weeks of the moderate carbohydrate diet relative to the traditional diet (34). A separate meta-analysis of single-arm trials estimated the change in HbA1c following the three single-arm low carbohydrate diet interventions (25, 31, 32) among 19 youth and 38 adults with type 1 diabetes. Pooled results demonstrated an HbA1c reduction of -0.63% (95%CI - 0.99, -0.27%) (i.e., -6.9 mmol/mol [95%CI –10.8, -3.0]) over 4.9 ± 0.96 months of a low carbohydrate diet (**Figure 1B**; **Supplemental Table 8**).

**Figure 1.**
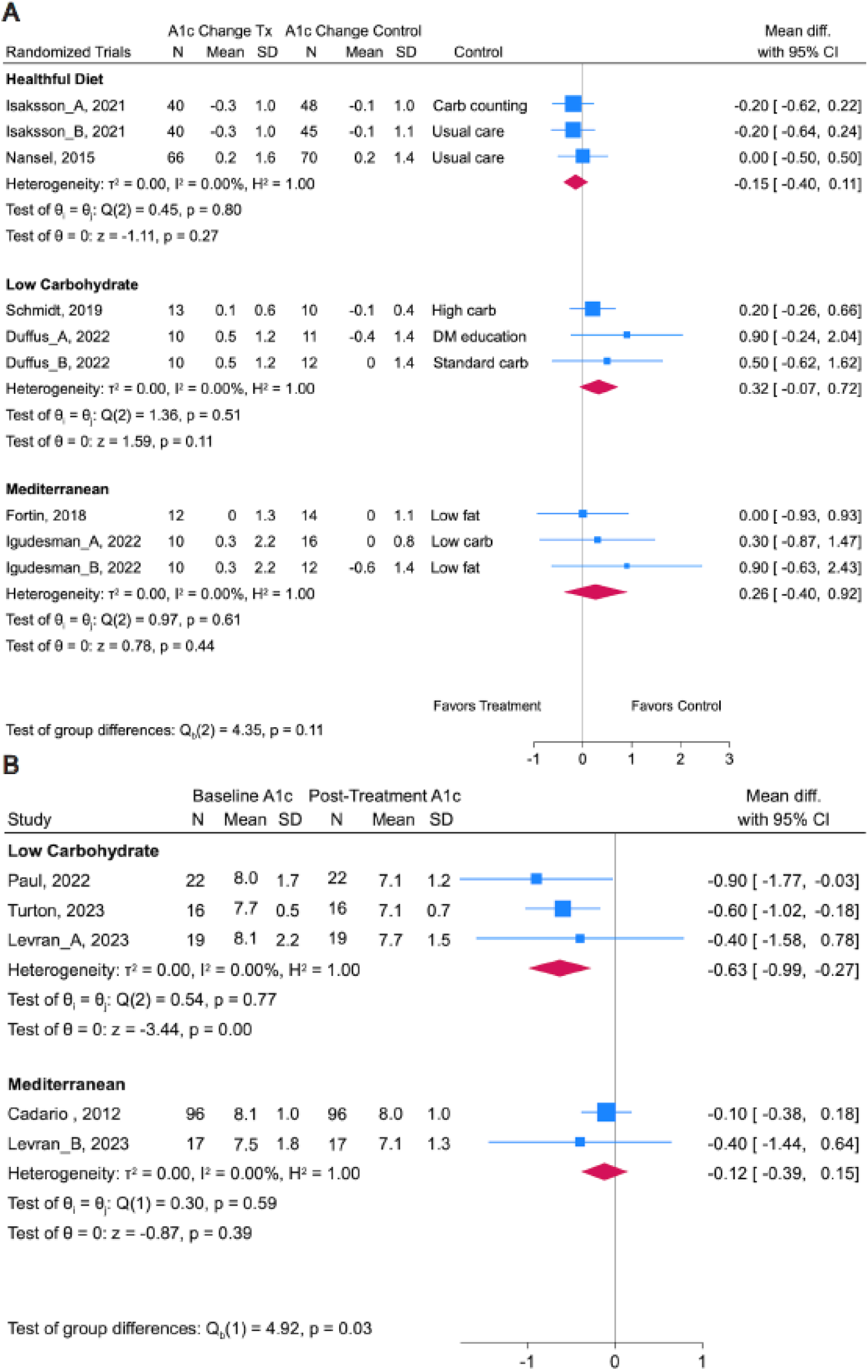
Forest plots showing pooled standard mean differences and 95%CIs from random effects regression models for randomized (**A**) and single-arm clinical trials (**B**) reporting HbA1c. The suffixes “_A” and “_B” were used to report separate effect estimates when more than one control arm was included. Tests for heterogeneity indicate whether effects varied across studies and include τ^2^ (i.e., Tau), I^2^, H^2^, and L_i_ (i.e., the Cochrane Q test). Degrees of freedom (n-1 studies) are shown in parentheses followed by the chi-square value and p-value for the Cochrane Q test. The test of L indicates whether the pooled result for each diet pattern and study type is different from the null. The test of group differences indicates whether the pooled results for randomized trials vary by diet pattern. Values for Nansel *et al.* estimated from visual presentation of results. HbA1c values for individual studies included in meta-analyses are reported in mmol/mol in **Supplemental Tables 7** and **8**. Values were converted from % to mmol/mol using ngsp.org/convert1.asp.

##### Mediterranean Diet Pattern

Two randomized (21, 27) and two single-arm trials (26, 33) investigated the effects of a eucaloric Mediterranean diet pattern on HbA1c. A meta-analysis of the randomized trials involving 52 adults with type 1 diabetes (duration 4.5 ± 2.1 months) found that a Mediterranean diet without caloric restriction had no differential effects on HbA1c relative to a hypocaloric low carbohydrate diet (15-20% calories from carbohydrates) (21), a hypocaloric moderate low fat diet, and a eucaloric low fat diet (both <30% calories from fat) (21, 27) (0.26% [95%CI –0.40, 0.92]; 2.8 mmol/mol [95%CI -4.4, 10.1]). Supporting the pooled results of the randomized trials in adults, a meta-analysis of the two single-arm trials implementing a eucaloric Mediterranean diet (duration 7.4 ± 1.9 months) in 123 youth with type 1 diabetes indicated no change in HbA1c pre- and post-treatment (0.12% [95%CI –0.39, 0.15]; 1.3 mmol/mol [95%CI –4.3, 1.6]).

##### Comparison of Dietary Patterns

The test for group differences across randomized trials indicated no differences in the effects of healthful, low carbohydrate, or Mediterranean diet patterns on HbA1c (p=0.11). However, there was a statistically significant difference across diet patterns among single-arm interventions (p=0.03), reflecting a pooled HbA1c reduction following low carbohydrate but not Mediterranean diets (no healthful diet patterns were included).

#### Impact of Dietary Patterns on Weight

##### Healthful Diet Pattern

Insufficient weight data from two randomized trials of healthful diet patterns (29, 35) rendered a meta-analysis of this diet pattern infeasible. Neither study found a differential effect of a healthful diet intervention relative to control (carbohydrate counting (35), routine care without a diet intervention (35), or attentional control matched for frequency of contacts (29)) on weight change over 12 (35) or 18 months (29) among 136 youth (29) and 133 adults (35) with type 1 diabetes.

##### Low Carbohydrate Diet Pattern

Two randomized crossover trials of low or moderate carbohydrate diets reported weight data (28, 34); however, a meta-analysis was not feasible as the moderate carbohydrate intervention did not report sufficient pre-intervention data(34). The moderate carbohydrate diet had no differential effect on weight relative to a traditional diet (50% energy from carbohydrates) in 50 adults with type 1 diabetes (0.0 kg difference in weight change [95%CI −0.4, 0.4 kg]). The other randomized trial lasting 3 months showed a ∼4 kg greater weight loss in 14 adults following a low versus high carbohydrate diet (28). A randomized low carbohydrate diet intervention found no difference in BMI change among 33 adolescents and young adults randomized to four months of a low carbohydrate diet, a standard carbohydrate diet, or general diabetes education, although the reported change in carbohydrate consumption was small, with only one participant adhering to recommended carbohydrate intake (30). A meta-analysis of two low carbohydrate single-arm trials involving 48 adults with type 1 diabetes corroborated the findings from the narratively synthesized randomized trials, showing no statistically significant change in weight over 4.3 ± 0.0 months (–2.01kg [95%CI –7.87, 3.85 kg]; **Figure 2B**). However, Levran *et al.* reported a –0.13 (IQR - 0.29, -0.02) reduction in BMI z-score in 19 youth and young adults with type 1 diabetes following a 6-month single-arm low carbohydrate diet intervention (25).

**Figure 2.**
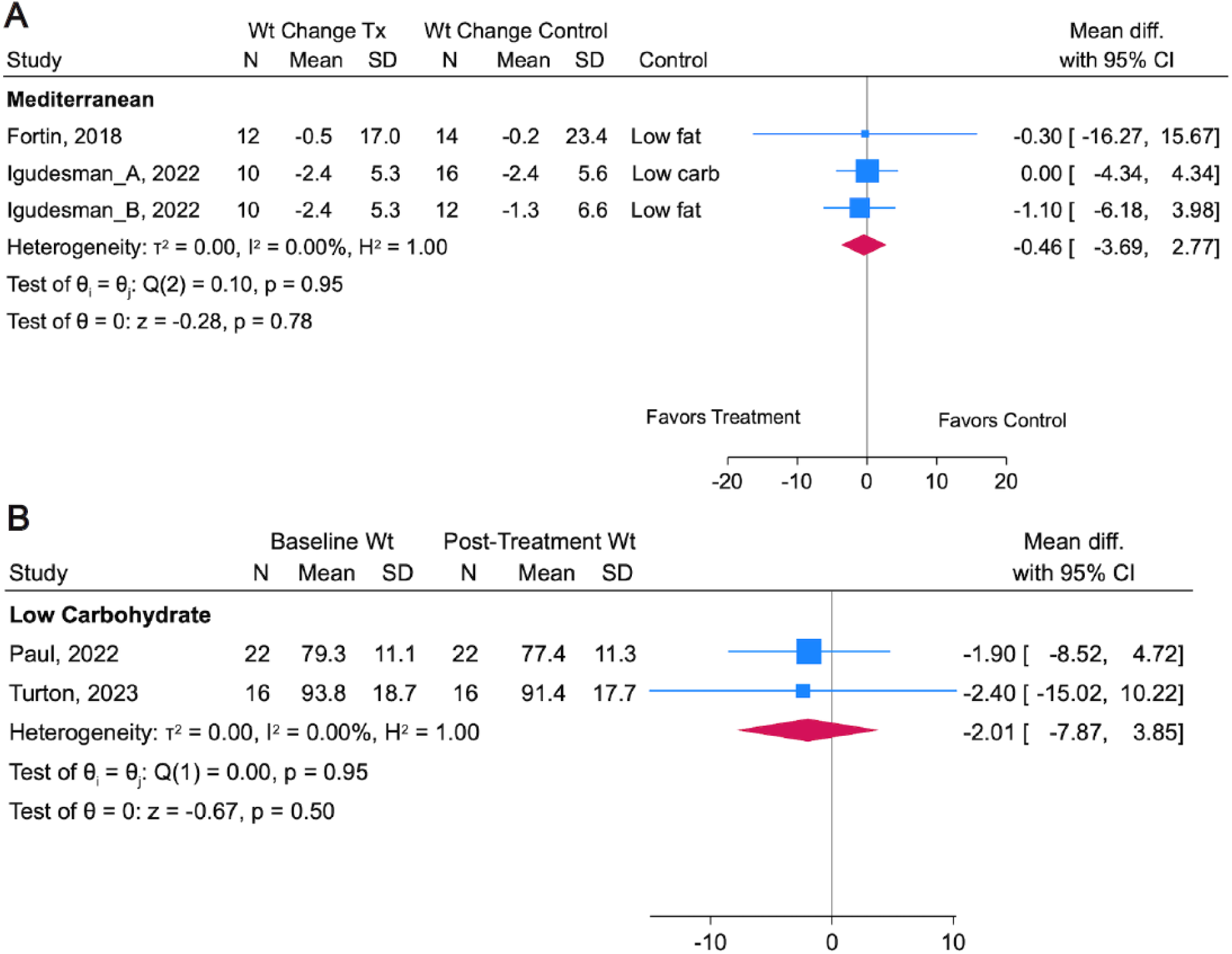
Forest plots showing pooled point estimates and 95%CIs from random effects regression models for randomized (**A**) and single-arm clinical trials (**B**) reporting weight (kg). The suffixes “_A” and “_B” were used to report separate effect estimates when more than one control arm was included. Tests for heterogeneity indicate whether effects varied across studies and include τ^2^ (i.e., Tau), I^2^, H^2^, and LJ_i_ (i.e., the Cochrane Q test). Degrees of freedom (n-1 studies) are shown in parentheses followed by the chi-square value and p-value for the Cochrane Q test. The test of LJ indicates whether the pooled result for each diet pattern and study type is different from the null.

##### Mediterranean Diet Pattern

The pooled results of a meta-analysis describing the effects of two randomized interventions of Mediterranean diets on weight change in 64 adults showed no difference relative to a eucaloric low fat diet (27), a hypocaloric low carbohydrate diet (21), or a hypocaloric low fat (21) (–0.46 kg [95%CI –3.69, 2.77 kg]). Supporting these results, two single-arm Mediterranean diet trials found no change in BMI z-score among youth and/or young adults with type 1 diabetes over six months (25, 33).

#### Impact of Low Fat Dietary Pattern on Hemoglobin A1c and Weight

One study involving 12 participants randomized to three months of a hypocaloric low fat diet found no differential change in HbA1c or weight relative to a hypocaloric low carbohydrate diet (n=16) or a eucaloric Mediterranean diet (n=10) (21).

#### Insulin dose

Most studies measured changes in insulin dose via self-report (32, 35) or did not disclose the method of measurement (25, 26, 31, 33, 34). One healthful diet study including 148 adults over 12 months found no difference in total daily insulin relative to carbohydrate counting or usual care (35). One (28) of three randomized trials (28, 30, 34) and all three single-arm trials (25, 31, 32) of low or moderate carbohydrate diets found 12-week reductions in bolus insulin (28, 31) or in total daily dose over 3-6 months in 20 adolescents (25) and 20 adults (**Supplemental Table 7**, **Supplemental Table 8**). Two of these studies found no change in basal insulin relative to a high carbohydrate diet control (28) or pre-post diet (31). Two single-arm Mediterranean diet studies reported changes in total daily insulin dose in adolescents with type 1 diabetes over 6 months, with one finding a 5-unit increase in total daily dose (33), and the other a trend towards a decrease in the insulin to weight ratio (26).

#### Heterogeneity

I^2^ values were 0 for all pooled analyses, indicating a lack of heterogeneity in results across trials included in meta-analyses. A select number of studies reported within-study variation in outcomes, which can only be summarized using narrative synthesis given insufficient data on outcomes of interest. One single-arm trial investigating a low carbohydrate diet in adults with type 1 diabetes compared the characteristics of responders (n=10) and non-responders (n=12), defined as a ≥10% or <10% relative reduction in HbA1c, respectively (31). Pre-intervention HbA1c was ∼2 percentage points higher in responders, whereas diet adherence and weight loss were not related to treatment response (31).

Two Mediterranean diet interventions reported subgroup analyses by sex (27, 33). Fortin *et al.* found no diet by sex interactions for HbA1c or weight when comparing the effects of a Mediterranean diet with a low fat diet in adults with T1D (27). The single-arm Mediterranean diet intervention in youth with type 1 diabetes found that HbA1c decreased by 0.2% (2.2 mmol/mol) over 6 months among pubertal boys, with no change appreciated in prepubertal boys or girls regardless of puberty status (33). In addition, BMI-SDS decreased by ∼0.2 units among prepubertal boys and in youth with overweight or obesity but increased among prepubertal girls (33).

Igudesman *et al.* reported participant characteristics associated with weight and glycemic outcomes irrespective of whether participants were randomized to a eucaloric Mediterranean diet, a hypocaloric low carbohydrate diet, or a hypocaloric low fat diet (21). The authors noted that non-Hispanic White participants had a 0.28 greater percentage point (95%CI 0.10, 1.23) HbA1c improvement than their counterparts from any other racial and ethnic group who comprised ∼34% of the sample, and that men had a 0.62 greater HbA1c percentage point reduction (95%CI 0.02, 1.2; i.e., 6.8 mmol/mol [95%CI 0.2, 13.1]) than women, after correction for baseline HbA1c (21). Women also lost 2.6 (95%CI 0.41, 4.9) fewer kilograms of weight than men, even after adjusting for baseline weight.

## Conclusions

While medical nutrition therapy is a crucial aspect of diabetes management, the present study highlights the dearth of evidence supporting specific nutrition guidelines for persons with type 1 diabetes in whom unique physiological and behavioral considerations (36) warrant dedicated nutrition research. In this systematic review and meta-analysis, we evaluated 12 studies that addressed the impact of dietary interventions on glycemia and weight in individuals with type 1 diabetes. Our pooled meta-analyses of randomized trials suggested no added benefit of any particular diet pattern on glycemia or weight relative to control interventions. Imprecision was graded as serious, raising the possibility that certain population subgroups may benefit more from specific diets than others. Changes in insulin dose were documented in some but not all studies and often measured via self-report; these data are critical for interpretation and should be rigorously measured in future studies.

Conversely, pooled results from single-arm interventions suggest reduced HbA1c with a low carbohydrate diet, albeit from a very low certainty body of evidence. The major caveat noted is that, in addition to lack of a control group, there was no adjustment made for time-varying confounders in any of the included trials, making the probability of confounding bias by changes in physical activity, caloric intake, and other variables likely. At least one single-arm low carbohydrate trial reported reductions in caloric intake (5) over time, which could explain changes in weight, insulin doses, and HbA1c independent of diet composition. Recognizing these limitations and without assessment of a low carbohydrate intervention in children younger than 12, cautionary guidance has been issued by the International Society for Pediatric and Adolescent Diabetes (ISPAD) and the American Academy of Pediatrics (37, 38) against adopting this diet in children with type 1 diabetes given a lack of long-term safety and efficacy data.

Meta-analyses of low certainty randomized trials testing the effects of generally healthful diet patterns or Mediterranean diets failed to show differential effects on HbA1c or weight. A meta-analysis of very low certainty single-arm Mediterranean diets showed analogous results for HbA1c. Nonetheless, wide 95%CIs indicated clinically meaningful heterogeneity that should be explored in future dietary trials of sufficient size and diversity to permit subgroup analysis. Despite lacking sufficient data to perform meta-analyses in sample subgroups to inform future precision nutrition guidelines, here, we present the first narrative synthesis of evidence in support of specific diets according to sex, weight status, pubertal status, and HbA1c, but the low quality of evidence and small number of studies precludes making tailored recommendations. Much of this preliminary evidence is derived from single-arm trials with a high risk of bias or from randomized trials with modest sample sizes, so studies with the explicit aim to study heterogeneity must be conducted.

### Precision Nutrition: Future Integration with Published Society Guidelines

The ADA and ISPAD recommend individualized medical nutrition therapy as an essential aspect of treatment for individuals living with type 1 diabetes. Despite the ADA’s emphasis on individualizing eating plans with a variety of foods that incorporate personal and cultural preferences, evidence for stratifying recommendations to optimize health within population subgroups is lacking (39, 40). The scant evidence base addressing diet approaches for managing overweight and obesity in type 1 diabetes (36) warrants particular attention. The ADA Standards of Medical Care focus exclusively on weight in the context of type 2 diabetes in the chapter titled Obesity and Weight Management (41). Our findings highlight that future research is urgently needed to rigorously examine the impact of dietary interventions on weight in those with type 1 diabetes, with particular attention to potential subgroups of interest.

Precision nutrition approaches must be vigilant not to widen disparities. This will require the purposive sampling of historically underrepresented groups in research to prevent algorithm bias, which is a consideration for artificial intelligence (AI) algorithms that are becoming commonplace in precision nutrition research. This may lead to suboptimal performance of AI-enabled clinical practice guidelines in these subgroups (42). Further, it is critical that researchers are cognizant of enrolling older adults with type 1 diabetes. A recent systematic review and summary of clinical practice guidelines addressing nutritional status, dietary intake, and eating behaviors in older adults with type 1 diabetes revealed substantial knowledge gaps in a group already at high risk for suboptimal health outcomes (43). Therefore, several factors need to be considered in trial design, including recruitment across the age spectrum of people with type 1 diabetes from diverse demographic backgrounds, regardless of baseline glycemia as those with an HbA1c substantially above target stand to receive the most benefit. Trial diversity—which was limited in the included studies—will ensure recommended dietary strategies take into account cultural food preferences and affordability and are (45) equitable and sustainable. One previous report did address the opportunity for healthful eating with food costs similar to that of less healthful diets (46).

### Strengths and Limitations

#### Strengths

Strengths of this work include a clear, focused research question, quantitative syntheses from stratified meta-analyses, a comprehensive literature search with a transparent, rigorous methodology, and detailed risk of bias and quality assessments. Studies enrolled participants across the lifespan and numerous geographic regions, which facilitates generalizability to a variety of target populations. All studies but one four-week intervention (34) were three months in duration or greater, so nearly all interventions were sufficiently long to observe changes in HbA1c and weight.

#### Limitations

Our review focused on assignment to an intervention rather than diet adherence. Most studies enrolled relatively few (<50) participants and lasted six months or less, making it difficult to evaluate generalizability and diet sustainability. Some studies were not included in the meta-analysis due to missing data and were instead narratively synthesized, which is less objective than quantitative synthesis even when done systematically (24). Although meta-analyses indicated a lack of heterogeneity between studies, variation in the control groups studied, the populations sampled, and intervention duration made pooled results challenging to interpret. Due to only one study being available for evaluation, the Inconsistency criterion for GRADE could not be assessed for randomized studies of low carbohydrate diets studying weight, nor for randomized trials of low fat interventions studying HbA1c and weight. Only one study explicitly focused on weight loss (21), which may in part explain minimal changes in weight in most other trials. While most studies included participants using an insulin pump, CGM, or both, only one study included those using low glucose suspend and predictive low-glucose suspend technology (28). This may limit generalizability given the potential for automated insulin delivery systems to liberalize dietary intake (e.g., by reducing fear of hypoglycemia) and modify the efficacy of medical nutrition therapy—gaps in knowledge that future studies should address. Studies that reported changes in insulin dose often did not disclose measurement methods or collected this information via self-report. Insulin dose data were not meta-analyzed. Changes in background and adjunctive therapies over time may have confounded the results of single-arm trials. Larger, long-term studies with deep participant phenotyping are needed to evaluate the feasibility and sustainability of AI-enabled diet recommendations and address longer-term impacts of nutritional intake on diabetes-related complications.

### Future Directions and Implications for Clinical Practice

While optimizing glycemia and weight is critical for cardiovascular disease mitigation, future reviews should evaluate the effects of diet patterns on comprehensive cardiovascular risk factors including hypertension and dyslipidemia as advocated by the ADA (44). CGM data should be collected in all future diet studies enrolling participants with type 1 diabetes to evaluate more granular effects of diet patterns on glycemia as well as the safety of diets—particularly those targeting weight loss through caloric restriction. Future studies should also investigate the effects of diet patterns on enteroendocrine hormone responses that impact postprandial glucose, gastric emptying, insulin needs, appetite, and body adiposity. Other deep phenotypic measures of human physiology and the gut microbiota should be investigated for their usefulness in training AI algorithms to optimize dietary recommendations (45). In addition to physiological measures, psychosocial outcomes are critical for preserving quality of life in type 1 diabetes, and experimental diets have been shown to have discordant effects on psychosocial outcomes and clinical endpoints in persons with type 1 diabetes (46). Future research is needed to guide individualized dietary approaches for subgroups with specific comorbidities (e.g., chronic kidney disease, celiac disease), and those taking adjunctive therapies (e.g., GLP-1 agonists, SGLT2-inhibitors). Weight loss studies should strive to measure body composition as minimizing lean body mass loss is important, particularly in older adult populations.

Controlled feeding studies such as those employed by the Nutrition for Precision Health Initiative can elucidate physiological responses to diet interventions independent of diet adherence and determine subpopulations that benefit more from specific diet approaches (47). A recent randomized crossover controlled feeding study published after we concluded the present review found that five weeks of outpatient low carbohydrate feeding (94.5 ± 4.7 g/day) increased time in target glucose range by ∼3 percentage points relative to a recommended carbohydrate diet (191 ± 19.2 g/day) in 34 youth with type 1 diabetes (47). Even with this level of dietary control, the 95%CI for change in time in range spanned 1 to 5 percentage points, indicating clinically significant heterogeneity between participants. Behavioral trials will be a vital complement to controlled feeding studies to evaluate longer-term feasibility. Rigorous assessments of diet adherence with food photography (48), doubly-labeled water (49) and novel dietary biomarkers should be employed (NCT05621863).

Our results support the emphasis of ADA and ISPAD guidelines on personalizing diet patterns to individual goals, preferences, cultures, and physiology. Further research is needed to determine whether nutrition guidelines should be tailored to specific population subgroups to reduce cardiovascular disease burden in persons with type 1 diabetes. Larger studies of longer duration and higher quality are needed to develop the evidence base that will guide AI-enabled precision health approaches, which hold the promise to optimize the clinical management of all individuals living with type 1 diabetes.

## Data Availability

Template data collection forms; data extracted from included studies; and analytic code will be made available upon reasonable request.

## Acknowledgments

The authors thank Thomas Mead for his peer-review of the search strategy.

## Funding and Assistance

This work was supported by the National Institutes of Health under K23 DK128560.

## Author Contributions and Guarantor Statement

All authors contributed to the study concept. DI, LMN, and AAG designed the search strategy, and AAG performed the search. DI and LMN take full responsibility for the work as a whole, including the study design, access to data, and the decision to submit and publish the manuscript. DI, LMN reviewed and analyzed the data, drafted the manuscript, prepared tables and figures, and finalized the manuscript. AAG, and CG contributed to the initial manuscript. CG performed statistical analyses. All authors reviewed and approved the final manuscript.

