## Supplemental Tables and Figures for "Dietary patterns for weight and glycemic management in persons with type 1 diabetes: a meta-analysis of clinical trials"

**Dietary patterns for weight and glycemic management in persons with type 1 diabetes: a precision nutrition-focused systematic review and meta-analysis of clinical trials**

Daria Igudesman^1^*, Laura Nally^2^*, Alyssa A. Grimshaw^3^, Craig Gunderson^2^, Elizabeth G. Considine^2^, Laura Jacobsen^4^, Mustafa Tosur^5^, Peter Gottlieb^6^, Irl Hirsch^7^, Lori Laffel^8^, Jennifer Sherr^2^, Chantal Mathieu^9^, Richard E. Pratley^1^

*These authors contributed equally:

^1^AdventHealth Translational Research Institute, Orlando, FL, USA

^2^Yale University School of Medicine, New Haven, CT, USA

^3^Harvey Cushing/John Hay Whitney Medical Library, Yale Unversity, New Haven, CT USA

^4^University of Florida School of Medicine, Gainesville, FL, USA

^5^Texas Children’s Hospital, Baylor College of Medicine, Houston, TX, USA

^6^Barbara Davis Center for Diabetes, University of Colorado School of Medicine, Aurora, CO, USA

^7^University of Washington School of Medicine, Seattle, WA, USA

^8^Joslin Diabetes Center, Harvard Medical School, Boston, MA, USA

^9^University Leuven, Leuven, Belgium

**Supplemental Table 1: PRISMA 2020 Main Reporting Guideline Checklists**

| **Topic** | **No.** | **Item** | **Location where item is reported** |
| --- | --- | --- | --- |
| **TITLE** |  |  |  |
| **Title** | 1 | Identify the report as a systematic review. | p. 1 |
| **ABSTRACT** |  |  |  |
| **Abstract** | 2 | See the PRISMA 2020 for Abstracts checklist | Suppl. |
| **INTRODUCTION** |  |  |  |
| **Rationale** | 3 | Describe the rationale for the review in the context of existing knowledge. | p. 8-9 |
| **Objectives** | 4 | Provide an explicit statement of the objective(s) or question(s) the review addresses. | p. 9 |
| **METHODS** |  |  |  |
| **Eligibility criteria** | 5 | Specify the inclusion and exclusion criteria for the review and how studies were grouped for the syntheses. | p. 10-11 |
| **Information sources** | 6 | Specify all databases, registers, websites, organisations, reference lists and other sources searched or consulted to identify studies. Specify the date when each source was last searched or consulted. | p. 10 |
| **Search strategy** | 7 | Present the full search strategies for all databases, registers and websites, including any filters and limits used. | Suppl. |
| **Selection process** | 8 | Specify the methods used to decide whether a study met the inclusion criteria of the review, including how many reviewers screened each record and each report retrieved, whether they worked independently, and if applicable, details of automation tools used in the process. | p. 11-12 |
| **Data collection process** | 9 | Specify the methods used to collect data from reports, including how many reviewers collected data from each report, whether they worked independently, any processes for obtaining or confirming data from study investigators, and if applicable, details of automation tools used in the process. | p. 12 |
| **Data items** | 10a | List and define all outcomes for which data were sought. Specify whether all results that were compatible with each outcome domain in each study were sought (e.g. for all measures, time points, analyses), and if not, the methods used to decide which results to collect. | p. 12 |
|  | 10b | List and define all other variables for which data were sought (e.g. participant and intervention characteristics, funding sources). Describe any assumptions made about any missing or unclear information. | p. 12 |
| **Study risk of bias assessment** | 11 | Specify the methods used to assess risk of bias in the included studies, including details of the tool(s) used, how many reviewers assessed each study and whether they worked independently, and if applicable, details of automation tools used in the process. | p. 12-13 |
| **Effect measures** | 12 | Specify for each outcome the effect measure(s) (e.g. risk ratio, mean difference) used in the synthesis or presentation of results. | p. 14 |
| **Synthesis methods** | 13a | Describe the processes used to decide which studies were eligible for each synthesis (e.g. tabulating the study intervention characteristics and comparing against the planned groups for each synthesis (item 5)). | p. 14-15 |
|  | 13b | Describe any methods required to prepare the data for presentation or synthesis, such as handling of missing summary statistics, or data conversions. | p. 14 |
|  | 13c | Describe any methods used to tabulate or visually display results of individual studies and syntheses. | p. 14 |
|  | 13d | Describe any methods used to synthesize results and provide a rationale for the choice(s). If meta-analysis was performed, describe the model(s), method(s) to identify the presence and extent of statistical heterogeneity, and software package(s) used. | p. 14 |
|  | 13e | Describe any methods used to explore possible causes of heterogeneity among study results (e.g. subgroup analysis, meta-regression). | p. 14-15 |
|  | 13f | Describe any sensitivity analyses conducted to assess robustness of the synthesized results. | N/A |
| **Reporting bias assessment** | 14 | Describe any methods used to assess risk of bias due to missing results in a synthesis (arising from reporting biases). | N/A |
| **Certainty assessment** | 15 | Describe any methods used to assess certainty (or confidence) in the body of evidence for an outcome. | p. 13-14 |
| **RESULTS** |  |  |  |
| **Study selection** | 16a | Describe the results of the search and selection process, from the number of records identified in the search to the number of studies included in the review, ideally using a flow diagram. | p. 15, Suppl. |
|  | 16b | Cite studies that might appear to meet the inclusion criteria, but which were excluded, and explain why they were excluded. | p. 15 |
| **Study characteristics** | 17 | Cite each included study and present its characteristics. | p. 14-15 |
| **Risk of bias in studies** | 18 | Present assessments of risk of bias for each included study. | p. 24, Suppl. |
| **Results of individual studies** | 19 | For all outcomes, present, for each study: (a) summary statistics for each group (where appropriate) and (b) an effect estimate and its precision (e.g. confidence/credible interval), ideally using structured tables or plots. | p. 16-22 |
| **Results of syntheses** | 20a | For each synthesis, briefly summarise the characteristics and risk of bias among contributing studies. | 13 |
|  | 20b | Present results of all statistical syntheses conducted. If meta-analysis was done, present for each the summary estimate and its precision (e.g. confidence/credible interval) and measures of statistical heterogeneity. If comparing groups, describe the direction of the effect. | 14-18 |
|  | 20c | Present results of all investigations of possible causes of heterogeneity among study results. | 19 |
|  | 20d | Present results of all sensitivity analyses conducted to assess the robustness of the synthesized results. | NA |
| **Reporting biases** | 21 | Present assessments of risk of bias due to missing results (arising from reporting biases) for each synthesis assessed. | 13 |
| **Certainty of evidence** | 22 | Present assessments of certainty (or confidence) in the body of evidence for each outcome assessed. | 13-14 |
| **DISCUSSION** |  |  |  |
| **Discussion** | 23a | Provide a general interpretation of the results in the context of other evidence. | 23, 26-28 |
|  | 23b | Discuss any limitations of the evidence included in the review. | 25-26 |
|  | 23c | Discuss any limitations of the review processes used. | 25-26 |
|  | 23d | Discuss implications of the results for practice, policy, and future research. | 26-28 |
| **OTHER INFORMATION** |  |  |  |
| **Registration and protocol** | 24a | Provide registration information for the review, including register name and registration number, or state that the review was not registered. | 2 |
|  | 24b | Indicate where the review protocol can be accessed, or state that a protocol was not prepared. | 2 |
|  | 24c | Describe and explain any amendments to information provided at registration or in the protocol. | NA |
| **Support** | 25 | Describe sources of financial or non-financial support for the review, and the role of the funders or sponsors in the review. | 28 |
| **Competing interests** | 26 | Declare any competing interests of review authors. | 28-30 |
| **Availability of data, code and other materials** | 27 | Report which of the following are publicly available and where they can be found: template data collection forms; data extracted from included studies; data used for all analyses; analytic code; any other materials used in the review. | 30 |

From: Page MJ, McKenzie JE, Bossuyt PM, Boutron I, Hoffmann TC, Mulrow CD, et al. The PRISMA 2020 statement: an updated guideline for reporting systematic reviews. MetaArXiv. 2020, September 14. DOI: 10.31222/osf.io/v7gm2. For more information, visit: www.prisma-statement.org

**Supplemental Table 2: PRISMA Abstract Reporting Checklist**

| **Topic** | **No.** | **Item** | **Reported?** |
| --- | --- | --- | --- |
| **TITLE** |  |  |  |
| **Title** | 1 | Identify the report as a systematic review. | Yes |
| **BACKGROUND** |  |  |  |
| **Objectives** | 2 | Provide an explicit statement of the main objective(s) or question(s) the review addresses. | Yes |
| **METHODS** |  |  |  |
| **Eligibility criteria** | 3 | Specify the inclusion and exclusion criteria for the review. | Yes |
| **Information sources** | 4 | Specify the information sources (e.g. databases, registers) used to identify studies and the date when each was last searched. | Yes |
| **Risk of bias** | 5 | Specify the methods used to assess risk of bias in the included studies. | Yes |
| **Synthesis of results** | 6 | Specify the methods used to present and synthesize results. | Yes |
| **RESULTS** |  |  |  |
| **Included studies** | 7 | Give the total number of included studies and participants and summarise relevant characteristics of studies. | Yes |
| **Synthesis of results** | 8 | Present results for main outcomes, preferably indicating the number of included studies and participants for each. If meta-analysis was done, report the summary estimate and confidence/credible interval. If comparing groups, indicate the direction of the effect (i.e. which group is favoured). | Yes |
| **DISCUSSION** |  |  |  |
| **Limitations of evidence** | 9 | Provide a brief summary of the limitations of the evidence included in the review (e.g. study risk of bias, inconsistency and imprecision). | Yes |
| **Interpretation** | 10 | Provide a general interpretation of the results and important implications. | Yes |
| **OTHER** |  |  |  |
| **Funding** | 11 | Specify the primary source of funding for the review. | Yes |
| **Registration** | 12 | Provide the register name and registration number. | Yes |

**Supplemental Table 3: Search Strategies**

**Ovid Embase**

1	exp insulin dependent diabetes mellitus/

2	((brittle or type-1 or type-i or type-I or type-one or type-1a or type-Ia or type 1-5 or double or insulin-depend* or juvenile or labile or ketoacidotic or early-onset or autoimmune or auto-immune or ketosis-prone or sudden-onset or immune-mediated or insulin-treated or fulminant or insulin-deficien*) adj3 diabet*).tw,kf.

3	(dm-1 or iddm or mckusick-22210 or T1DM or t1d or lada).ti,ab.

4	1 or 2 or 3

5	Dietary pattern/

6	exp atkins diet/ or healthy diet/ or exp ketogenic diet/ or low carbohydrate diet/ or exp low fiber diet/ or exp mediterranean diet/ or paleolithic diet/ or exp vegetarian diet/

7	exp Healthy Eating Index/

8	exp dash diet/ or low fat diet/ or low glycemic index diet/

9	exp carbohydrate intake/

10	((health* or quality) adj3 (diet* or eating* or food*)).tw,kf.

11	((diet* or eating*) adj3 (habit* or pattern*)).tw,kf.

12	((Mediterranean or low-fat* or low-carb or low-carbohydrate* or moderate-carbohydrate* or carbohydrate restricted or carbohydrate poor or low-sodium or plant-based or keto* or paleo* or MIND or MAD or low GI or low glycemic or reduced GI or HPLC or HP-LC) adj5 (meal* or diet*)).tw,kf.

13	(MedDiet or DASH or look-AHEAD or HEI or Dietary-Approach*-to-Stop-Hypertension or Atkins or LGIT or vegan or vegetarian* or pescatarian*).tw,kf.

14	or/5-13

15	4 and 14

16	limit 15 to yr="2011 -Current"

17	exp animal/

18	exp animal/ and exp human/

19	17 not 18

20	16 not 19

21	limit 20 to conference abstracts

22	20 not 21

**Ovid MEDLINE(R) ALL**

1	exp Diabetes Mellitus, Type 1/

2	((brittle or type-1 or type-i or type-I or type-one or type-1a or type-Ia or type 1-5 or double or insulin-depend* or juvenile or labile or ketoacidotic or early-onset or autoimmune or auto-immune or ketosis-prone or sudden-onset or immune-mediated or insulin-treated or fulminant or insulin-deficien*) adj3 diabet*).tw,kf.

3	(dm-1 or iddm or mckusick-22210 or T1DM or t1d or lada).ti,ab.

4	1 or 2 or 3

5	exp diet, carbohydrate-restricted/ or exp diet, fat-restricted/ or diet, mediterranean/ or diet, paleolithic/ or diet, reducing/ or diet, sodium-restricted/ or exp diet, vegetarian/ or exp diet, western/ or dietary approaches to stop hypertension/ or diet, healthy/

6	exp Dietary Carbohydrates/

7	((health* or quality) adj3 (diet* or eating* or food*)).tw,kf.

8	((diet* or eating*) adj3 (habit* or pattern*)).tw,kf.

9	((Mediterranean or low-fat* or low-carb or low-carbohydrate* or moderate-carbohydrate* or carbohydrate restricted or carbohydrate poor or low-sodium or plant-based or keto* or paleo* or MIND or MAD or low GI or low glycemic or reduced GI or HPLC or HP-LC) adj5 (meal* or diet*)).tw,kf.

10	(MedDiet or DASH or look-AHEAD or HEI or Dietary-Approach*-to-Stop-Hypertension or Atkins or LGIT or vegan or vegetarian* or pescatarian*).tw,kf.

11	5 or 6 or 7 or 8 or 9 or 10

12	4 and 11

13	limit 12 to yr="2011 -Current"

14	exp animals/

15	exp animals/ and exp humans/

16	14 not 15

17	13 not 16

**AMED (Allied and Complementary Medicine)**

1	diabetes mellitus type 1/

2	((brittle or type-1 or type-i or type-I or type-one or type-1a or type-Ia or type 1-5 or double or insulin-depend* or juvenile or labile or ketoacidotic or early-onset or autoimmune or auto-immune or ketosis-prone or sudden-onset or immune-mediated or insulin-treated or fulminant or insulin-deficien*) adj3 diabet*).mp.

3	(dm-1 or iddm or mckusick-22210 or T1DM or t1d or lada).ti,ab.

4	1 or 2 or 3

5	diet fads/ or diet mediterranean/ or diet vegetarian/ or exp infant nutrition/

6	dietary carbohydrates/

7	((health* or quality) adj3 (diet* or eating* or food*)).mp.

8	((diet* or eating*) adj3 (habit* or pattern*)).mp.

9	((Mediterranean or low-fat* or low-carb or low-carbohydrate* or moderate-carbohydrate* or carbohydrate restricted or carbohydrate poor or low-sodium or plant-based or keto* or paleo* or MIND or MAD or low GI or low glycemic or reduced GI or HPLC or HP-LC) adj5 (meal* or diet*)).mp.

10	(MedDiet or DASH or look-AHEAD or HEI or Dietary-Approach*-to-Stop-Hypertension or Atkins or LGIT or vegan or vegetarian* or pescatarian*).mp.

11	or/5-10

12	4 and 11

13	limit 12 to yr="2011 -Current"

**Web of Science Core Collection**

#1 TS=((brittle or type-1 or type-i or type-I or type-one or type-1a or type-Ia or double or insulin-depend* or juvenile or labile or ketoacidotic or early-onset or autoimmune or auto-immune or ketosis-prone or sudden-onset or immune-mediated or insulin-treated or fulminant or insulin-deficien*) near/3 diabet*) or TS=(dm-1 or iddm or mckusick-22210 or T1DM or t1d or lada)

#2 TS=((health* or quality) near/3 (diet* or eating* or food*)) or TS=((diet* or eating*) near/3 (habit* or pattern*)) or TS=((Mediterranean or low-fat* or low-carb or low-carbohydrate* or moderate-carbohydrate* or "carbohydrate restricted" or "carbohydrate poor" or low-sodium or plant-based or keto* or paleo* or MIND or MAD or "low GI" or "low glycemic" or "reduced GI" or HPLC or HP-LC) near/5 (meal* or diet*)) or TS=(MedDiet or DASH or look-AHEAD or HEI or Dietary-Approach*-to-Stop-Hypertension or Atkins or LGIT or vegan or vegetarian* or pescatarian*)

#3 #1 and #2

#4 #1 AND #2 and 2023 or 2021 or 2022 or 2020 or 2019 or 2018 or 2017 or 2016 or 2015 or 2014 or 2013 or 2012 or 2011 (Publication Years)

The Core Collection included in this review includes:

1.	Science Citation Index Expanded (1900 - Data Searched)

2.	Social Sciences Citation Index (1900 - Date Searched)

3.	Art & Humanities - (1975 - Date Searched)

4.	Conference Proceedings Citation Index - Science (1991 - Date Searched)

5.	Conference Proceedings Citation Index - Social Sciences and Humanities (1991 - Date Searched)

6.	Book Citation Index - Science (2005 - Date Searched)

7.	Book Citation Index - Social Sciences and Humanities ( 2005 - Date Searched)

8.	Emerging Source Citation Index - (2018 - Date Searched)

9.	Current Chemical Reactions (1985 - Date Searched)

10.	Index Chemicus (1993 - Date Searched)

**Cochrane Library**

#1 ((brittle or type-1 or type-i or type-I or type-one or type-1a or type-Ia or double or insulin-depend* or juvenile or labile or ketoacidotic or early-onset or autoimmune or auto-immune or ketosis-prone or sudden-onset or immune-mediated or insulin-treated or fulminant or insulin-deficien*) near/3 diabet*):ti,ab or (dm-1 or iddm or mckusick-22210 or T1DM or t1d or lada):ti,ab

#2 ((health* or quality) near/3 (diet* or eating* or food*)):ti,ab or ((diet* or eating*) near/3 (habit* or pattern*)):ti,ab or ((Mediterranean or low-fat* or low-carb or low-carbohydrate* or moderate-carbohydrate* or "carbohydrate restricted" or "carbohydrate poor" or low-sodium or plant-based or keto* or paleo* or MIND or MAD or "low GI" or "low glycemic" or "reduced GI" or HPLC or HP-LC) near/5 (meal* or diet*)):ti,ab or (MedDiet or DASH or look-AHEAD or HEI or Dietary-Approach-to-Stop-Hypertension or Atkins or LGIT or vegan or vegetarian* or pescatarian*):ti,ab

#3 #1 and #2

#4 limit to 2011 to 2023

The Cochrane Library database includes:

1.	Cochrane Database of Systematic Reviews

2.	Cochrane Central Register of Controlled Trials

3.	Cochrane Clinical Answers

**CINAHL**

S1 TI ( ((brittle or type-1 or type-i or type-I or type-one or type-1a or type-Ia or type 1-5 or double or insulin-depend* or juvenile or labile or ketoacidotic or early-onset or autoimmune or auto-immune or ketosis-prone or sudden-onset or immune-mediated or insulin-treated or fulminant or insulin-deficien*) N3 diabet*) ) OR AB ( ((brittle or type-1 or type-i or type-I or type-one or type-1a or type-Ia or type 1-5 or double or insulin-depend* or juvenile or labile or ketoacidotic or early-onset or autoimmune or auto-immune or ketosis-prone or sudden-onset or immune-mediated or insulin-treated or fulminant or insulin-deficien*) N3 diabet*) ) OR TI ( (dm-1 or iddm or mckusick-22210 or T1DM or t1d or lada) ) OR AB ( (dm-1 or iddm or mckusick-22210 or T1DM or t1d or lada) )

S2 (TI ( ((low or moderate or restricted or restrictive or poor) N3 (fat or carb* or sodium or glycemic or GI) N5 (diet* or meal*) ) ) OR AB ( ((low or moderate or restricted or restrictive or poor) N3 (fat or carb* or sodium or glycemic or GI) N5 (diet* or meal*) ) ) OR TI ( ((Mediterranean or plant-based or keto* or paleo*) N5 (meal* or diet*)) ) OR AB ( ((Mediterranean or plant-based or keto* or paleo*) N5 (meal* or diet*)) ) OR TI ( (MedDiet or DASH or look-AHEAD or HEI or Dietary-Approach*-to-Stop-Hypertension or Atkins or LGIT or vegan or vegetarian* or pescatarian*) ) OR AB ( (MedDiet or DASH or look-AHEAD or HEI or Dietary-Approach*-to-Stop-Hypertension or Atkins or LGIT or vegan or vegetarian* or pescatarian*) ) OR TI ( ((health* or quality) N3 (diet* or eating* or food*)) ) OR AB ( ((health* or quality) N3 (diet* or eating* or food*)) ) OR TI ( ((diet* or eating*) N3 (habit* or pattern*)) ) OR AB ( ((diet* or eating*) N3 (habit* or pattern*)) ))

S3 S1 and S2 Limited to 2011-2023

**Google Scholar (via Harzing’s Publish or Perish Edition: 8.7.4245.8399)**

type 1 diabetes dietary patterns

**Supplemental Table 4: Inclusion and Exclusion Criteria**

**Inclusion Criteria**

- Randomized and non-randomized clinical trials
- Trials ≥4 weeks in duration
- Youth or adults with type 1 diabetes aged ≥2 years enrolled
- Diabetes duration ≥6 months
- ≥10 participants per intervention arm, or 10 participants in total for randomized crossover trials.
- At least one of the co-primary review outcomes of hemoglobin A1c (HbA1c) or weight must have been reported.
- Published in a peer-reviewed journal between January 2011 and the date of the final search (June 13, 2024). Studies were published in English; studies published in another language were excluded during the abstract screening phase. Studies were grouped by study design (randomized or non-randomized) and diet pattern (low carbohydrate, Mediterranean, low fat, and general healthful eating).

**Exclusion Criteria**

- Observational studies, editorials/letters, opinions, reviews, theory, and non-human studies
- Studies that did not enroll participants with type 1 diabetes or did not clearly report results separately for this clinical population
- Studies that were restricted to participants with type 1 diabetes and co-occurring chronic kidney disease or celiac disease

**Supplemental Table 5.** **Synthesis without Meta-Analysis Checklist**

| **SWiM reporting item** | **Item description** | **Page in manuscript where item is reported** |
| --- | --- | --- |
| **Methods** | | |
| **1 Grouping studies for synthesis** | 1a) Provide a description of, and rationale for, the groups used in the synthesis (eg, groupings of populations, interventions, outcomes, study design) | We grouped studies by diet pattern (low or moderate carbohydrate, Mediterranean, low fat, and general healthful eating patterns) and design (randomized and single-arm; p.10). Trials that could not be meta-analyzed and were therefore narratively described included one randomized moderate carbohydrate diet for the outcome of HbA1c (Isaksson 2024, p.11). Two randomized healthful food based approaches (Isaksson, Nansel) did not report sufficient weight data for a meta analysis and the results are narratively synthesized on p. 14. Two randomized low or moderate carbohydrate diets reported weight but not in sufficient detail for a meta-analysis (Schmidt et al, Isaksson et al 2024), and a third reported BMI but not weight (Duffus et al); these three trials are narratively synthesized on p. 15. Two single-arm Mediterranean diet interventions reported BMI metrics and not weight and are narratively synthesized on p.16. Subgroup analyses were not meta-analyzed due to insufficient data and are narratively described on pp. 17-18. |
|  | 1b) Detail and provide rationale for any changes made subsequent to the protocol in the groups used in the synthesis | N/A |
| **2 Describe the standardised metric and transformation methods used** | Describe the standardised metric for each outcome. Explain why the metric(s) was chosen and describe any methods used to transform the intervention effects, as reported in the study, to the standardised metric, citing any methodological guidance consulted | Given that changes in the continuous variables of HbA1c and weight were meta-analyzed, pooled effects were reported in terms of mean difference in change in HbA1c (%) or weight (kg), as described on p. 10-11. |
| **3 Describe the synthesis methods** | Describe and justify the methods used to synthesise the effects for each outcome when it was not possible to undertake a meta-analysis of effect estimates | The results of individual studies were described narratively when <2 studies within each diet pattern and study design type (randomized and single-arm) had available data for a given outcome, and therefore could not be meta-analyzed. This is described on p. 11. |
| **4 Criteria used to prioritise results for summary and synthesis** | Where applicable, provide the criteria used, with supporting justification, to select the particular studies, or a particular study, for the main synthesis or to draw conclusions from the synthesis (eg, based on study design, risk of bias assessments, directness in relation to the review question) | All studies with available data were used to draw conclusions—both those that were included and were not included in meta-analyses. Pooled results are described first, followed by supporting data from individual studies that could not be meta-analyzed. Accordingly, risk of bias and GRADE assessments were completed for all included studies whether or not they were meta-analyzed, as described on p. 11. |
| **5 Investigation of heterogeneity in reported effects** | State the method(s) used to examine heterogeneity in reported effects when it was not possible to undertake a meta-analysis of effect estimates and its extensions to investigate heterogeneity | On p. 11 we note that the I^2^ metric was used to estimate between-study heterogeneity for each meta analysis. We narratively synthesized within-study heterogeneity in intervention effects within sociodemographic and clinical strata, as described on p. 11. |
| **6 Certainty of evidence** | Describe the methods used to assess the certainty of the synthesis findings | On p. 10 we described methods used to assess certainty of findings for both meta-analyses (i.e., 95% CIs associated with pooled estimates) and studies that could not be meta-analyzed (i.e., measures of dispersion as reported by individual study authors). Additionally, all included studies, whether meta-analyzed or not, were evaluated using the GRADE framework, as described on p. 10. |
| **7 Data presentation methods** | Describe the graphical and tabular methods used to present the effects (eg, tables, forest plots, harvest plots) | Description of forest plots used to display results and grouping of results described on p. 11. |
|  | Specify key study characteristics (eg, study design, risk of bias) used to order the studies, in the text and any tables or graphs, clearly referencing the studies included |  |
| **Results** | | |
| **8 Reporting results** | For each comparison and outcome, provide a description of the synthesised findings and the certainty of the findings. Describe the result in language that is consistent with the question the synthesis addresses, and indicate which studies contribute to the synthesis | Point estimates and 95%CIs from pooled results, as well as point estimates and 95%CIs or other measures of dispersion from individual studies that could not be meta-analyzed, are described on pp. 12-18. The latter includes results stratified by participant subgroups, as reported by study authors. |
| **Discussion** | | |
| **9 Limitations of the synthesis** | Report the limitations of the synthesis methods used and/or the groupings used in the synthesis and how these affect the conclusions that can be drawn in relation to the original review question | Pp. 22-23 |

**Supplemental Table 6: Excluded Studies**

| **First Authors Last Name** | **Year** | **Title** | **Journal** | **Reason for Exclusion** |
| --- | --- | --- | --- | --- |
| Ahola | 2018 | Association between diet and measures of arterial stiffness in type 1 diabetes - Focus on dietary patterns and macronutrient substitutions | Nutr Metab Cardiovasc Dis | Wrong intervention |
| Ahola | 2019 | Dietary carbohydrate intake and cardio-metabolic risk factors in type 1 diabetes | Diabetes Res Clin Pract | Wrong study design |
| Ahola | 2016 | Dietary patterns are associated with various vascular health markers and complications in type 1 diabetes | J Diabetes Complications | Wrong intervention |
| Antoniotti | 2022 | Adherence to the Mediterranean Diet Is Associated with Better Metabolic Features in Youths with Type 1 Diabetes | Nutrients | Wrong study design |
| Ayano-Takahara | 2015 | Carbohydrate intake is associated with time spent in the euglycemic range in patients with type 1 diabetes | Journal of Diabetes Investigation | Wrong study design |
| Balk | 2016 | Association of diet and lifestyle with glycated haemoglobin in type 1 diabetes participants in the EURODIAB prospective complications study | Eur J Clin Nutr | Wrong intervention |
| Bansal | 2013 | Dietary quality in adolescents with type 1 diabetes | Diabetes Care | Wrong study design |
| Barnes | 2013 | Change in DASH diet score and cardiovascular risk factors in youth with type 1 and type 2 diabetes mellitus: The SEARCH for Diabetes in Youth Study | Nutr Diabetes | Wrong study design |
| Bodur | 2021 | The Relationship Between Diet Quality of Adolescents with Type 1 Diabetes and Nutritional Status and Biochemical Parameters | Erciyes Med. J. | Wrong study design |
| Calella | 2023 | Lifestyle and physical fitness in adolescents with type 1 diabetes and obesity | Heliyon | Wrong study design |
| Colberg | 2021 | Physical Activity, Dietary Patterns, and Glycemic Management in Active Individuals with Type 1 Diabetes: An Online Survey | Int J Environ Res Public Health | Wrong study design |
| Dluzniak-Golaska | 2019 | Analysis of the diet quality and dietary habits of children and adolescents with type 1 diabetes | Diabetes Metab Syndr Obes | Wrong intervention |
| Dominguez-Riscart | 2022 | Adherence to Mediterranean Diet Is Associated With Better Glycemic Control in Children With Type 1 Diabetes: A Cross-Sectional Study | Front | Wrong study design |
| Ghaemi | 2021 | Effects of a Mediterranean diet on the development of diabetic complications: A longitudinal study from the nationwide diabetes report of the National Program for Prevention and Control of Diabetes (NPPCD 2016-2020) | Maturitas | Wrong study design |
| Gingras | 2015 | Association between cardiometabolic profile and dietary characteristics among adults with type 1 diabetes mellitus | J Acad Nutr Diet | Wrong study design |
| Grabia | 2021 | Adherence to Mediterranean Diet and Selected Lifestyle Elements among Young Women with Type 1 Diabetes Mellitus from Northeast Poland: A Case-Control COVID-19 Survey | Nutrients | Wrong outcomes |
| Kelleher | 2023 | Associations among permissive parenting, family mealtime behaviors, nutrition, and blood glucose in children with type 1 diabetes | Child. Health Care | Wrong intervention |
| Kleiner | 2022 | Safety and Efficacy of Eucaloric Very Low-Carb Diet (EVLCD) in Type 1 Diabetes: A One-Year Real-Life Retrospective Experience | Nutrients | Wrong study design |
| Koca | 2022 | Nutritional Habits, Compliance with Healthy Diet and Insulin Therapy, Depression and Family Functionality in Children with Type 1 Diabetes Mellitus during the Covid-19 Pandemic Period | Acta Endocrinol | Wrong study design |
| Krebs | 2016 | A randomised trial of the feasibility of a low carbohydrate diet vs standard carbohydrate counting in adults with type 1 diabetes taking body weight into account | Asia Pac J Clin Nutr | Sample size <10 per group |
| Krzyzowska | 2019 | Assessment of selected food intake frequency in patients with type 1 diabetes treated with personal insulin pumps | Rocz Panstw Zakl Hig | Wrong study design |
| Lehmann | 2020 | Lower Daily Carbohydrate Intake Is Associated With Improved Glycemic Control in Adults With Type 1 Diabetes Using a Hybrid Closed-Loop System | Diabetes Care | Wrong study design |
| Lennerz | 2018 | Management of Type 1 Diabetes With a Very Low-Carbohydrate Diet | Pediatrics | Wrong study design |
| Leow | 2018 | The glycaemic benefits of a very-low-carbohydrate ketogenic diet in adults with Type 1 diabetes mellitus may be opposed by increased hypoglycaemia risk and dyslipidaemia | Diabetic Med. | Observational study lacking analyses that link exposure and outcome |
| Leroux | 2015 | In adult patients with type 1 diabetes healthy lifestyle associates with a better cardiometabolic profile | Nutr Metab Cardiovasc Dis | Wrong intervention |
| Liese | 2020 | Association between diet quality indices and arterial stiffness in youth with type 1 diabetes: SEARCH for Diabetes in Youth Nutrition Ancillary Study | J Diabetes Complications | Wrong outcomes |
| Liese | 2011 | Association of DASH diet with cardiovascular risk factors in youth with diabetes mellitus: the SEARCH for Diabetes in Youth study | Circulation | Wrong study design |
| Marquard | 2011 | A prospective clinical pilot-trial comparing the effect of an optimized mixed diet versus a flexible low-glycemic index diet on nutrient intake and HbA<sub>1c</sub> levels in children with type 1 diabetes | J. Pediatr. Endocrinol. Metab. | Sample size <10 per group |
| Meissner | 2014 | Carbohydrate intake in relation to BMI, HbA1c and lipid profile in children and adolescents with type 1 diabetes | Clin Nutr | Wrong intervention |
| Muntis | 2023 | A High Protein Diet Is Associated with Improved Glycemic Control Following Exercise among Adolescents with Type 1 Diabetes | Nutrients | Wrong study design |
| Nansel | 2016 | Greater diet quality is associated with more optimal glycemic control in a longitudinal study of youth with type 1 diabetes | Am J Clin Nutr | Wrong study design |
| Nansel | 2015 | Little variation in diet cost across wide ranges of overall dietary quality among youth with type 1 diabetes | J Acad Nutr Diet | Wrong study design |
| Nansel | 2012 | Multiple indicators of poor diet quality in children and adolescents with type 1 diabetes are associated with higher body mass index percentile but not glycemic control | J Acad Nutr Diet | Wrong study design |
| Neuman | 2021 | Low-Carbohydrate Diet among Children with Type 1 Diabetes: A Multi-Center Study | Nutrients | Wrong study design |
| Nielsen | 2012 | Low carbohydrate diet in type 1 diabetes, long-term improvement and adherence: A clinical audit | Diabetol Metab Syndr | Wrong study design |
| Patton | 2013 | Dietary adherence and mealtime behaviors in young children with type 1 diabetes on intensive insulin therapy | J Acad Nutr Diet | Wrong study design |
| Powers | 2016 | Eating Patterns and Food Intake of Persons with Type 1 Diabetes | Diabetes | Duplicate |
| Powers | 2018 | Eating patterns and food intake of persons with type 1 diabetes within the T1D exchange | Diabetes Res Clin Pract | Wrong study design |
| Queiroz | 2012 | Influence of the glycemic index and glycemic load of the diet in the glycemic control of diabetic children and teenagers | Nutr Hosp | Wrong study design |
| Sanjeevi | 2018 | Cardiovascular Biomarkers in Association with Dietary Intake in a Longitudinal Study of Youth with Type 1 Diabetes | Nutrients | Wrong study design |
| Zhong | 2016 | Association of adherence to a Mediterranean diet with glycemic control and cardiovascular risk factors in youth with type I diabetes: the SEARCH Nutrition Ancillary Study | Eur J Clin Nutr | Wrong study design |

**Supplemental Table 7. HbA1c Values in mmol/mol for randomized trials shown in forest plots**

| **Study** | **Duration** | **Diet Pattern: Active Intervention** | **Control Intervention** | **Active Intervention Mean HbA1c change (mmol/mol)** | **Active Intervention SD of HbA1c change (mmol/mol)** | **Control Intervention Mean HbA1c change (mmol/mol)** | **Control Intervention SD of HbA1c change (mmol/mol)** | **Mean (95%CI) difference (mmol/mol)** |
| --- | --- | --- | --- | --- | --- | --- | --- | --- |
| Isaksson_A 2021 | 12 months | Healthful diet | Carbohydrate counting | -3.3 | 10.9 | -1.1 | 10.9 | -2.2 (-6.8, 2.4) |
| Isaksson_B 2021 | 12 months | Healthful diet | Usual care | -3.3 | 10.9 | -1.1 | 12.0 | -2.2 (-7.0, 2.6) |
| Nansel 2015 | 18 months | Healthful diet | Usual care | 2.2 | 17.5 | 2.2 | 15.3 | 0.0 (-5.5, 5.5)* |
| Schmidt 2019 | 12-week diet w/12-week washout | Low carbohydrate | High carbohydrate | 1.1 | 6.6 | -1.1 | 4.4 | 2.2 (-2.8, 7.2) |
| Duffus_A 2022 | 12 weeks | Low carbohydrate | Diabetes education | 5.5 | 13.1 | -4.4 | 15.3 | 9.8 (-2.6, 22.3) |
| Duffus_B 2022 | 12 weeks | Low carbohydrate | Standard carbohydrate | 5.5 | 13.1 | 0.0 | 15.3 | 5.5 (-6.8, 17.7) |
| Isaksson 2024** | 12 weeks | Moderate carbohydrate | Traditional | -- | -- | -- | -- | -- |
| Fortin 2018 | 6 months | Mediterranean | Low fat | 0.0 | 14.2 | 0.0 | 12.0 | 0.0 (-10.2, 10.2) |
| Igudesman_A 2022 | 3 months | Mediterranean | Hypocaloric low carbohydrate | 3.3 | 24.0 | 0.0 | 8.7 | 3.3 (-9.5, 16.1) |
| Igudesman_B 2022 | 3 months | Mediterranean | Hypocaloric low fat | 3.3 | 24.0 | -6.6 | 15.3 | 9.8 (-6.9, 15.6) |

*In Isaksson 2021, participants self-reported prandial insulin doses over 4 days. Values reported in Schmidt et al. are from 12 weeks of insulin pump downloads. Numeric values not reported in Duffus *et al.* but were collected from pump downloads in two-week increments throughout the study. Method of measuring changes in insulin dose not reported in Isaksson 2024.

**Insufficient HbA1c data were reported in Isaksson 2024 for inclusion in pooled analysis, so HbA1c values are not reported here.

**Supplemental Table 8. HbA1c Values in mmol/mol for non-randomized trials shown in forest plots**

| **Study** | **Duration** | **Diet Pattern: Active Intervention** | **Baseline Mean HbA1c change (mmol/mol)** | **Baseline SD of HbA1c change (mmol/mol)** | **Post-Treatment Mean HbA1c change (mmol/mol)** | **Post-Treatment SD of HbA1c change (mmol/mol)** | **Mean (95%CI) difference (mmol/mol)** |
| --- | --- | --- | --- | --- | --- | --- | --- |
| Paul 2022 | 12 weeks | Low carbohydrate | 64.0 | 18.6 | 54.0 | 13.1 | -9.8 (-19.3, 0.3) |
| Turton 2023 | 12 weeks | Low carbohydrate | 61.0 | 5.5 | 54.0 | 7.7 | -6.6 (-11.1, -2.0) |
| Levran_A 2023 | 6 months | Low carbohydrate | 65.0 | 24.0 | 61.0 | 16.4 | -4.4 (-17.3, 8.5) |
| Cadario 2012 | 6 months | Mediterranean | 65.0 | 10.9 | 64.0 | 10.9 | -1.1 (-4.2, 2.0) |
| Levran_B 2023 | 6 months | Mediterranean | 58.0 | 19.7 | 54.0 | 14.2 | -4.4 (-15.7, 7.0) |

HbA1c: hemoglobin A1c, CI: confidence interval, TDI: total daily insulin, IQR: interquartile range

Pooled results of meta-analyses are displayed in mmol/mol in manuscript text.

*****In Paul *et al,* Cadario *et al*, Levran_A, and Levran_B, the method of measuring changes in insulin dose was not stated. In Turton *et al*, a 3-day self-reported insulin log measured total daily insulin, or by pump downloads among those managing diabetes with an insulin pump.

**Supplemental Table 9. Changes in Total Daily Insulin Dose Alongside HbA1c and Weight: Randomized Trials**

| **Study** | **Duration** | **Diet Pattern: Active Intervention** | **Control Intervention** | **Mean difference weight (95% CI), kg*** | **Mean difference HbA1c (95% CI), %** | **Total Daily Insulin (TDI) dose**** |
| --- | --- | --- | --- | --- | --- | --- |
| Isaksson_A 2021 | 12 months | Healthful diet | Carbohydrate counting | -- | -0.20 (-0.62, 0.22) | Difference in TDI: 0.03 ± 0.02 U/kg body weight (p=0.16) |
| Isaksson_B 2021 | 12 months | Healthful diet | Usual care | -- | 0.20 (-0.64, 0.24) | Difference in TDI: 0.03 ± 0.02 U/kg body weight (p=0.08) |
| Nansel 2015 | 18 months | Healthful diet | Usual care | -- | 0.00 (0.40, 0.11) | Not reported |
| Schmidt 2019 | 12-week diet w/12-week washout | Low carbohydrate | High carbohydrate | -- | 0.20 (-0.26, 0.66) | TDI low carb: 33.6 ± 8.1 U,  TDI high carb: 43.2 ± 11.0 U (p<0.001)  Total daily basal low carb: 18.5 ± 5.4 U,  Total daily basal high carb: 17.3 ± 5.4 U (p=0.1)  Total daily bolus low carb: 15.1 ± 4.4 U  Total daily bolus high carb: 25.9 ± 7.3 U, (p<0.0001) |
| Duffus_A 2022 | 12 weeks | Low carbohydrate | Diabetes education | -- | 0.90 (-0.24, 2.04) | No group differences in insulin dose per unit body weight |
| Duffus_B 2022 | 12 weeks | Low carbohydrate | Standard carbohydrate | -- | 0.50 (-0.62, 1.62) | No group differences in insulin dose per unit body weight |
| Isaksson 2024*** | 12 weeks | Moderate carbohydrate | Traditional | -- | -- | Difference in TDI −3.3 U (−7.5 to 0.9), p=0.12 |
| Fortin 2018 | 6 months | Mediterranean | Low fat | -0.30 (-16.27, 15.67) | 0.00 (-0.93, 0.93) | Not reported |
| Igudesman_A 2022 | 3 months | Mediterranean | Hypocaloric low carbohydrate | 0.00 (-6.18, 3.98) | 0.30 (-0.87, 1.47) | Not reported |
| Igudesman_B 2022 | 3 months | Mediterranean | Hypocaloric low fat | -0.46 (-3.69, 2.77) | 0.90 (-0.63, 2.43) | Not reported |

HbA1c: hemoglobin A1c, CI: confidence interval, TDI: total daily insulin

*Pooled analyses for weight were only completed for Fortin 2018 and Igudesman 2022.

**In Isaksson 2021, participants self-reported prandial insulin doses over 4 days. Values reported in Schmidt et al. are from 12 weeks of insulin pump downloads. Numeric values not reported in Duffus *et al.* but were collected from pump downloads in two-week increments throughout the study. Method of measuring changes in insulin dose not reported in Isaksson 2024.

***Insufficient HbA1c data were reported in Isaksson 2024 for inclusion in pooled analysis, so HbA1c values are not reported here.

**Supplemental Table 10. Changes in Total Daily Insulin Alongside HbA1c and Weight: Single-Arm Trials**

| **Study** | **Duration** | **Diet Pattern: Active Intervention** | **Mean difference weight (95% CI), kg*** | **Mean difference HbA1c (95%CI), %** | **Change in total daily insulin dose**** |
| --- | --- | --- | --- | --- | --- |
| Paul 2022 | 12 weeks | Low carbohydrate | -1.90 (-8.52, 4.72) | -0.90 (-1.77, -0.03) | -7.0 U (95% CI:- 4.9 to -9.0, p<0.001) |
| Turton 2023 | 12 weeks | Low carbohydrate | -2.40 (-15.02, 10.22) | -0.60 (-1.02, -0.18) | 16 ± 11 U, p<0.001) |
| Levran_A 2023 | 6 months | Low carbohydrate | -- | -0.40 (-1.58, 0.78) | -0.12 U/kg (IQR -0.18, -0.04), p=0.006 |
| Cadario 2012 | 6 months | Mediterranean | -- | -0.10 (-0.38, 0.18) | Increase from 37.3 ± 1.3 U to 42.4 ± 1.6 U (p<0.0001) |
| Levran_B 2023 | 6 months | Mediterranean | -- | -0.40, (-1.44, 0.64) | Change in TDI from 0.76 U/kg (0.64–0.97) to 0.72 U/kg (0.61–0.89), p=0.067 |

**Supplemental Table 11. Certainty of evidence assessed by Grading of Recommendations Assessment, Development, and Evaluation (GRADE).**

| **Outcome** | **Diet Pattern** | **Type of Study** | **No. of studies** | **Study design** | **Inconsistency** | **Indirectness** | **Imprecision** | **Publication bias** | **Certainty of Evidence** |
| --- | --- | --- | --- | --- | --- | --- | --- | --- | --- |
| **HbA1c** | LCD, MCD | RT | 3 | Serious | Not serious | Serious | Serious | Not serious | Very Low* |
|  | LCD | SA | 3 | Very Serious | Not serious | Serious | Serious | Not serious | Very Low^†^ |
|  | Mediterranean | RT | 2 | Not serious | Not serious | Serious | Serious | Not serious | Low^‡^ |
|  | Mediterranean | SA | 2 | Very Serious | Not serious | Serious | Serious | Not serious | Very Low^†^ |
|  | Low Fat | RT | 1 | Not serious | N/A | Serious | Serious | Not serious | Low^‡^ |
|  | Healthful diet | RT | 2 | Not serious | Not serious | Serious | Not serious | Not serious | Moderate^§^ |
| **Weight** | LCD, MCD | RT | 3 | Serious | Serious | Serious | Not serious | Not serious | Very Low^\|\|^ |
|  | LCD | SA | 2 | Very Serious | Not Serious | Serious | Not serious | Not serious | Very Low^¶^ |
|  | Mediterranean | RT | 2 | Not serious | Not serious | Serious | Serious | Not serious | Low^‡^ |
|  | Low Fat | RT | 1 | Not serious | N/A | Serious | Not serious | Not serious | Moderate^§^ |
|  | Healthful diet | RT | 1 | Not serious | N/A | Serious | Not serious | Not serious | Moderate^§^ |

Abbreviations: LCD: low carbohydrate diet, MCD: moderate carbohydrate diet, RT: randomized trial, SA: single arm

*downgrade 1 for study design (high risk of bias), 1 for indirectness (lack of representativeness), 1 for imprecision

^†^downgrade by 2 for study design (high risk of bias), 1 for indirectness (lack of representativeness), 1 for imprecision

^‡^ downgrade by 1 for indirectness (lack of representativeness), 1 for imprecision

^§^ downgrade 1 for indirectness (lack of representativeness)

^||^ downgrade by 1 for study design (high risk of bias), 1 for inconsistency, 1 for indirectness (lack of representativeness)

^¶^ downgrade by 2 for study design (high risk of bias), 1 for indirectness

**Supplemental text to accompany supplemental Table 11**

The strength of the evidence was graded as being very low certainty for both the three randomized (1-3) and three non-randomized trials (4-6) of low or moderate carbohydrate trials. This was true for both co-primary outcomes of HbA1c and weight. Each of the randomized low or moderate carbohydrate diet trials was deemed to have a high risk of bias in one domain for the outcome of HbA1c (1-3). For two out of three studies, this was due to missing data (1, 3), whereas for the third study, this was due to insufficient washout between study arms for the supplemental crossover domain (2).

In contrast to the randomized low carbohydrate trials, the two randomized Mediterranean diet trials were *not* found to have any serious limitations in the Risk of Bias GRADE domain for the outcomes of HbA1c and weight but were rather downgraded for indirectness and imprecision (7, 8). The two single-arm Mediterranean diet trials investigating HbA1c change were graded as providing very low certainty evidence for their effects on HbA1c (9, 10). Finally, the strength of evidence for the two randomized trials evaluating the effects of general healthful eating patterns on HbA1c (11, 12) and the sole healthful diet intervention to study effects on weight (11) was graded as moderate certainty. This was because these studies were downgraded only for indirectness.

**Supplemental Table 12.** Population, intervention, comparator, outcomes, and study designs (PICOS) framework

| **Criteria** | **Description** |
| --- | --- |
| **Population** | Children or adults with type 1 diabetes aged ≥2 years with diabetes duration of ≥6 months. Older adults consuming modified foods (i.e., pre-chewed, thickened, etc.) will be excluded. |
| **Intervention** | The diet exposure will be focused on diet patterns including low or moderate carbohydrate (or low glycemic index, Atkins, or ketogenic), low fat, Mediterranean (or MIND), DASH (or low-sodium), vegetarian, vegan, plant-based, paleolithic, and adherence to food-based guidelines such as the US Dietary Guidelines (i.e., Healthy Eating Index score). |
| **Comparators** | “Control” diet or other diet comparator. There may not be a comparator if the study is single-arm (i.e., pre-post), in which baseline values are used for comparison. and analyzes the diet variable continuously (i.e., per unit increase or decrease in adherence score). |
| **Outcomes** | - Hemoglobin A1c - Weight (kg) |
| **Study designs** | Randomized controlled trials, pilot randomized trials, non-randomized single-arm trials |

**Supplemental Figure 1.** PRISMA Flowchart

**Identification of studies via other methods**

**Identification of studies via databases and registers**

Records removed *before screening*:

Duplicates or Retrieved in Database Searching (n = 152)

Records identified from:

Citation searching (n = 494)

Records removed *before screening*:

- Duplicate records removed (n = 1588)
- Records marked as ineligible by automation tools (n = 0)
- Records removed for other reasons (n = 0)

Records identified from:

- Databases (n = 4637)
- Registers (n = 0)

**Identification**

Records excluded

(n = 342)

Records screened

(n = 342)

Records screened

(n = 3049)

Records excluded

(n = 2996)

Reports not retrieved

(n = 0)

Reports not retrieved

(n = 0)

Reports sought for retrieval

(n = 0)

Reports sought for retrieval

(n = 53)

**Screening**

Reports excluded: (n = 41)

- Wrong study design (n = 28)
- Wrong intervention (n = 7)
- Sample size < 10 (n = 2)
- Wrong outcomes (n = 2)
- Duplicate (n = 1)
- No analysis between exposure and outcome (n = 1)

Reports excluded: (n = 0)

Reports assessed for eligibility

(n = 0)

Reports assessed for eligibility

(n = 53)

Studies included in review

(n = 12)

**Included**

*Adapted from:* Page MJ, McKenzie JE, Bossuyt PM, Boutron I, Hoffmann TC, Mulrow CD, et al. The PRISMA 2020 statement: an updated guideline for reporting systematic reviews. BMJ 2021;372: n71. doi: 10.1136/bmj. n71. For more information, visit: <http://www.prisma-statement.org/>

**Supplemental Figure 2.**

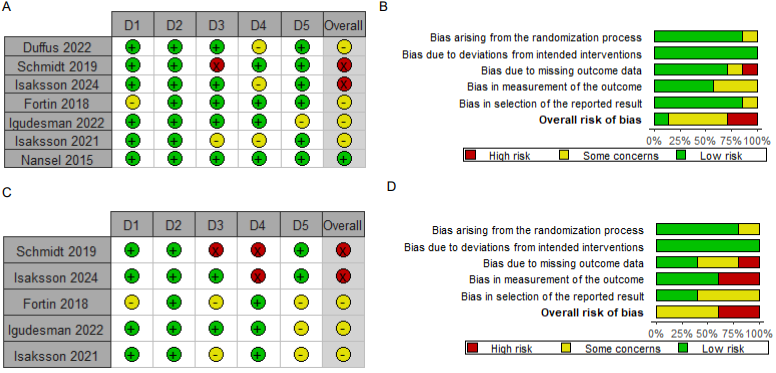

Panels A and B show risk of bias assessments for randomized trials using the RoB2 tool for HbA1c, whereas panels C and D are for weight. The supplemental crossover domain is not shown but was deemed to be high risk for Isaksson 2024 for the outcome of HbA1c due to insufficient washout time, resulting in an overall high risk of bias (A). Domains 1-5 (i.e., D1 – D5) shown in panels A and C are described in order in panels B and D. D1: bias arising from the randomization process; D2: Bias due to deviations from intended interventions; D3: Bias due to missing outcome data; D4: Bias in measurement of the outcome; D5: Bias in selection of the reported result.

**Supplemental Figure 3.**

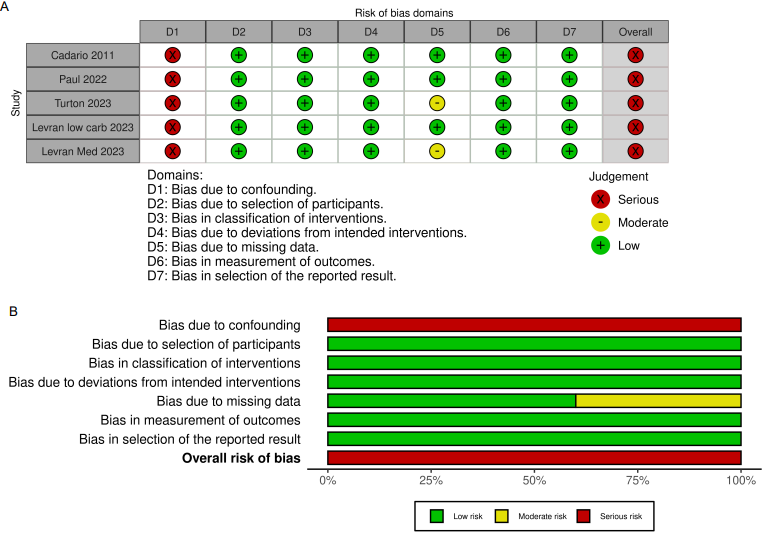

Traffic light (A) and bar plot (B) showing risk of bias assessments from the ROBINS-I tool for non-randomized trials reporting HbA1c.

**Supplemental Figure 4.**

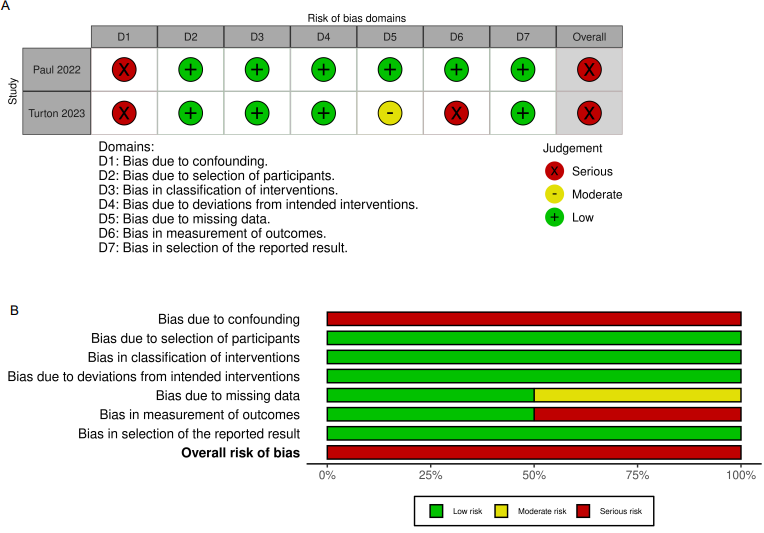

Traffic light (A) and bar plot (B) showing risk of bias assessments from the ROBINS-I tool for non-randomized trials reporting weight.
